# Extreme differences in SARS-CoV-2 viral loads among respiratory specimen types during presumed pre-infectious and infectious periods

**DOI:** 10.1101/2022.07.13.22277113

**Authors:** Alexander Viloria Winnett, Reid Akana, Natasha Shelby, Hannah Davich, Saharai Caldera, Taikun Yamada, John Raymond B. Reyna, Anna E. Romano, Alyssa M. Carter, Mi Kyung Kim, Matt Thomson, Colten Tognazzini, Matthew Feaster, Ying-Ying Goh, Yap Ching Chew, Rustem F. Ismagilov

**Affiliations:** California Institute of Technology, Pasadena, CA, USA; Pangea Laboratory LLC, Tustin, CA, USA; Zymo Research Corp., Irvine, CA, USA; Pasadena Public Health Department, Pasadena, CA, USA

## Abstract

SARS-CoV-2 viral load measurements from a single specimen type are used to establish diagnostic strategies, interpret clinical-trial results for vaccines and therapeutics, model viral transmission, and understand virus-host interactions. However, measurements from a single specimen type are implicitly assumed to be representative of other specimen types. We quantified viral-load timecourses from individuals who began daily self-sampling of saliva, anterior nares (nasal), and oropharyngeal (throat) swabs before or at the incidence of infection with the Omicron variant. Viral loads in different specimen types from the same person at the same timepoint exhibited extreme differences, up to 10^9^ copies/mL. These differences were not due to variation in sample self-collection, which was consistent. For most individuals, longitudinal viral-load timecourses in different specimen types did not correlate. Throat-swab and saliva viral loads began to rise up to 7 days earlier than nasal-swab viral loads in most individuals, leading to very low clinical sensitivity of nasal swabs during the first days of infection. Individuals frequently exhibited presumably infectious viral loads in one specimen type while viral loads were low or undetectable in other specimen types. Therefore, defining an individual as infectious based on assessment of a single specimen type underestimates the infectious period, and overestimates the ability of that specimen type to detect infectious individuals. For diagnostic COVID-19 testing, these three single specimen types have low clinical sensitivity, whereas a combined throat-nasal swab, and assays with high analytical sensitivity, were inferred to have significantly better clinical sensitivity to detect presumed pre-infectious and infectious individuals.

**Significance Statement:** In a longitudinal study of SARS-CoV-2 Omicron viral loads in three paired specimen types (saliva, anterior-nares swabs, and oropharyngeal swabs), we found extreme differences among paired specimen types collected from a person at the same timepoint, and that viral loads in different specimen types from the same person often do not correlate throughout infection. Individuals often exhibited high, presumably infectious viral loads in oral specimen types before nasal viral loads remained low or even undetectable. Combination oropharyngeal-nasal swabs were inferred to have superior clinical sensitivity to detect infected and infectious individuals. This demonstrates that single specimen type reference standard tests for SARS-CoV-2, such as in clinical trials or diagnostics evaluations may miss infected and even infectious individuals.

## INTRODUCTION

Measurements of viral load in respiratory infections are used to establish diagnostic strategies, interpret results of clinical trials of vaccines and therapeutics, model viral transmission, and understand virus-host interactions. But how viral loads change across multiple specimen types early in SARS-CoV-2 infection is not well understood. Specifically in the context of diagnostics, as new SARS-CoV-2 variants-of-concern (and new respiratory viruses) emerge with different viral kinetics(1), it is imperative to continually re-evaluate testing strategies (including specimen type and test analytical sensitivity) for detecting pre-infectious and infectious individuals. Early detection can reduce transmission within communities(2, 3) and the global spread of new variants, and enable earlier initiation of treatment resulting in better outcomes(4-6).

Selecting testing strategies to achieve detection in the pre-infectious and infectious periods of new SARS-CoV-2 variants or new viruses requires filling two critical knowledge gaps: [1] Which respiratory specimen type accumulates virus first? [2] What is the appropriate test analytical sensitivity to detect accumulation of virus in the pre-infectious and infectious stages? These two gaps must be filled in parallel.

Commonly, an individual’s infection is described by the viral load sampled from a single specimen type, which is appropriate when there is one principal specimen type (e.g., HIV in blood plasma). However, some respiratory infections, including SARS-CoV-2, can infect multiple respiratory sampling sites(7-9), thus viral loads could potentially differ between sampling sites throughout infection. Beyond diagnostics, many studies continue to use samples from a single specimen type to analyze viral infections, implicitly assuming that viral loads are the same across all specimen types. Data comparing viral load trajectories in multiple sample types are needed to evaluate this assumption.

Nasopharyngeal swabs have been the gold standard for SARS-CoV-2 detection but are poorly tolerated and challenging for serial sampling and self-collection. Many alternate specimen types are now widely used. Some of these are suitable for routine testing, and are approved for self-collection (e.g., saliva, anterior-nares (nasal) swabs, and oropharyngeal (throat) swabs) in some countries. Cross-sectional studies comparing paired specimen types from the same person have found that cycle threshold (Ct, a semi-quantitative proxy for viral load) values can differ significantly between specimen types and the clinical sensitivity of different specimen types is not equivalent(10).

In some cases, viral loads in one specimen type are low or even absent while viral loads in another type may be high(11-13). Nasal swabs (including those used for rapid antigen testing) continue to be the dominant specimen type used in the U.S., often for workplace screenings and at-home testing. However, several studies(14-17), news media(18), and social media posts have speculated that in Omicron infections, viral load accumulates in oral specimens before the nasal cavity. Formal investigations of specimen types from single timepoints and cross-sectional studies have been contradictory, so rigorous comparisons starting from the incidence of infection are needed.

The second knowledge gap is the analytical sensitivity needed for reliable detection of pre-infectious and infectious individuals. The assay analytical sensitivity is described by the limit of detection (LOD); generally, the LOD of an assay describes its ability to detect and quantify target at or above a certain concentration in that specimen type with >95% probability(19). Assays with high LODs (low analytical sensitivity) require a high concentration of virus to reliably yield positive results, whereas assays with low LODs (high analytical sensitivity) can reliably detect much lower concentrations of virus. For example, in early SARS-CoV-2 variants, some studies showed that saliva accumulated virus earlier than nasal swabs, but at low levels(13, 20, 21); therefore, detecting SARS-CoV-2 early in saliva required a high-analytical-sensitivity (i.e., low LOD) assay(13, 22). However, low-analytical-sensitivity tests (including rapid antigen tests) have been increasingly manufactured, authorized, and used globally(23, 24). Which of these tests can detect pre-infectious and infectious individuals requires quantitative, longitudinal measurements of viral concentration in multiple specimen types starting from the incidence of infection.

Early detection, in the pre-infectious period, is ideal to prompt infection-control practices (e.g., isolation) before transmission occurs, and detection during the infectious period is critical to minimize outbreaks. Replication-competent (i.e., infectious) virus has been recovered from saliva(9), oropharyngeal swabs(25), and nasal swabs(26), but it is both impractical and infeasible to perform viral culture on each positive specimen to determine if a person is infectious. However, studies that performed both culture and RT-qPCR found that low Ct values (high viral loads) are associated with infectious virus. Specific viral loads likely to be infectious for each specimen type have not been established(27), partly because Ct values are not comparable across assays(28, 29) and culture methods differ. However, as a general reference, viral loads of >10^4^–10^7^ RNA copies/mL are associated with the presence of replication-competent virus in culture(16, 30-40), and these values have been used in outbreak simulations(35, 39, 41-43). The enormous range (>4 orders of magnitude) in observed viral loads that correspond with infectiousness emphasizes why quantitative measurements of viral loads in different specimen types are needed to make robust predictions about tests that will detect the pre-infectious and infectious periods.

The assumption made early in the COVID-19 pandemic that viral load always rises rapidly from undetectable to likely infectious(44), has been challenged by numerous longitudinal studies of viral load in different specimen types that show early SARS-CoV-2 viral loads can rise slowly over days(13, 16, 17, 20, 26, 45-48), not hours. These finds are encouraging because this longer window can provide more time to identify and isolate pre-infectious individuals. However, making use of this opportunity by selecting an optimal diagnostic test requires a thorough understanding of how viral load changes in each specimen type early in infection for each variant. Similarly, to reliably detect an infectious person, an infectious specimen must be tested with an assay that has an LOD below the infectious viral load for that specimen type. However, many authorized COVID-19 tests (including rapid antigen tests) have LODs well above the range of reported infectious viral loads(49, 50).

Filling the two critical and inter-related knowledge gaps about specimen type and assay LOD requires high-frequency quantification of viral loads, rather than semi-quantitative Ct values, in multiple specimen types starting from the incidence of infection, not after a positive test or after symptom onset, as is commonly done. Moreover, quantification must be performed with a high-analytical-sensitivity assay to capture low viral loads in the first days of detectable infection. It is challenging to acquire such data. Individuals at high risk of infection must be prospectively enrolled prior to detectable infection and tested longitudinally with high-frequency in multiple paired specimen types.

To our knowledge, four studies have reported longitudinal viral-load timecourses in multiple, paired specimen types from early infection. A university study(26) captured daily saliva and nasal-swab samples for 2 weeks from 60 individuals, only 3 of whom were negative for SARS-CoV-2 upon enrollment. In our prior study, we captured twice-daily viral-load timecourses from 72 individuals for 2 weeks(51), 7 of whom were negative upon enrollment(13). In 6 of 7 individuals, we inferred from viral-load quantifications that a high-analytical-sensitivity saliva assay would detect infection earlier than a low-analytical-sensitivity nasal-swab test. **I**n a SARS-CoV-2 human challenge study(16), 10 of 18 infected participants had detectable virus by PCR in throat swabs at least one day prior to nasal swabs, and replication-competent virus was recovered from throat swabs before nasal swabs in at least 12 of 18 participants. However, participants in these three studies were infected with pre-Omicron variants. One longitudinal study(14) analyzed viral loads in three major specimen types (saliva, nasal swabs, and throat swabs) over time in Omicron. However, daily measurements in all three sample types were captured for only two individuals, both of whom were already positive upon enrollment. Thus, the rise and fall of Omicron viral loads in different specimen types from the incidence of infection has not been characterized, despite these data being necessary to define the appropriate test analytical sensitivity and specimen type to best detect pre-infectious and infectious individuals.

Here, we measured and analyzed the viral-load timecourses of the Omicron variant in three specimen types appropriate for self-sampling (saliva, nasal swabs, and throat swabs) by individuals starting at or before the incidence of infection as part of a household transmission study in Southern California. We then utilized these data to determine which specimen type and analytical sensitivity would yield the most reliable detection of pre-infectious and infectious individuals. A separate paper reports the results of daily rapid antigen testing in this study(52).

## MATERIALS AND METHODS

### Study Design

This case-ascertained study of household transmission was conducted in the greater Los Angeles County area between November 23, 2021, and March 1, 2022. This study was approved under Caltech IRB #20-1026. All adult participants provided written informed consent; all minor participants provided verbal assent accompanied by written permission from a legal guardian. Children ages 8-17 years old additionally provided written assent. Additional details can be found in the Supplemental Information.

A total of 228 participants from 56 households were enrolled in the study; 90 of whom tested positive for SARS-CoV-2 infection during enrollment (**Fig 1**). We limited our analyses to 14 individuals (**Table S1, Fig 2**) who enrolled in the study at or before the incidence of acute SARS-CoV-2 infection. To be included in the cohort, a participant must have had at least one specimen type with viral loads below quantification upon enrollment, followed by positivity and quantifiable viral loads in all three specimen types.

**Figure 1.**
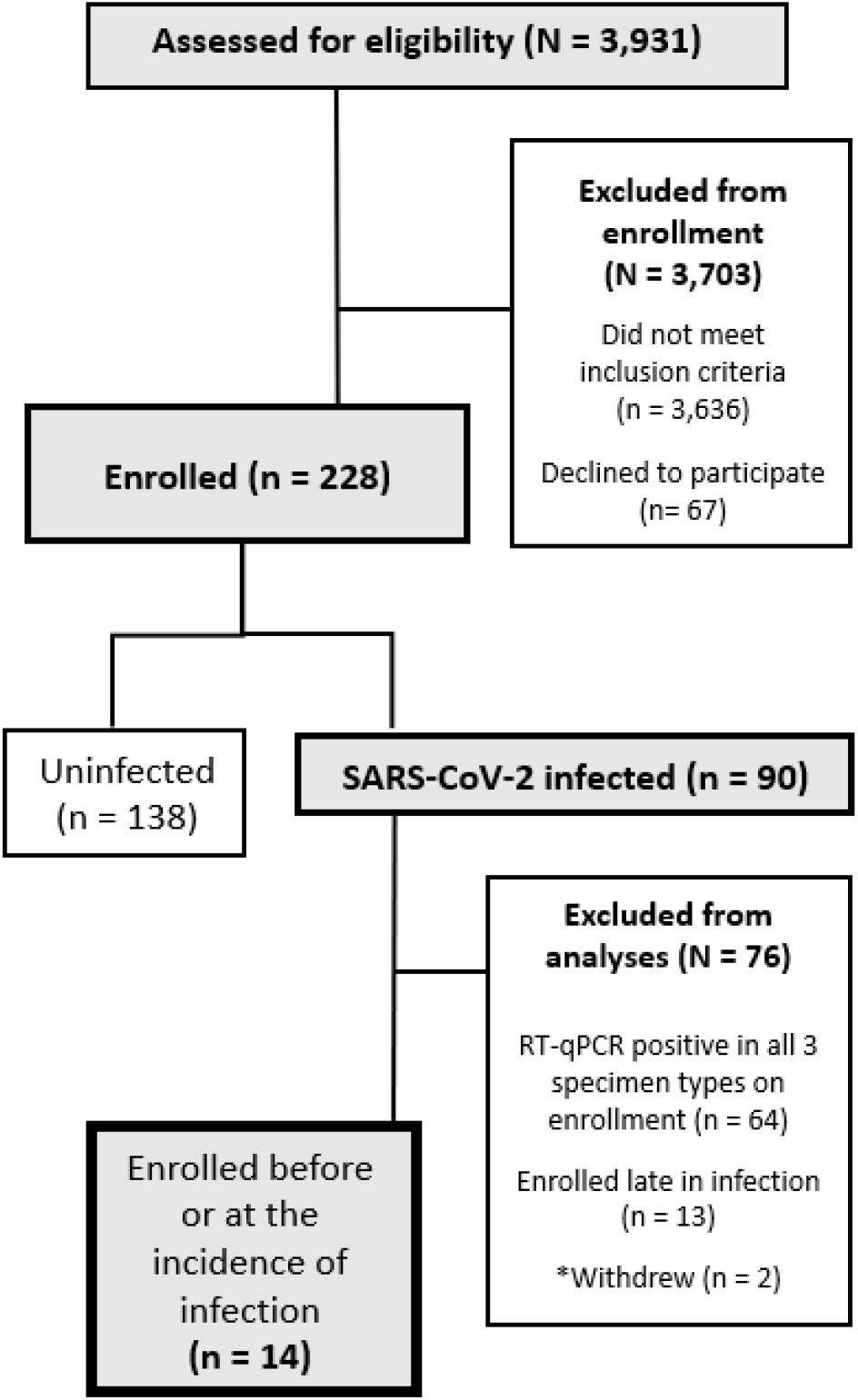
A CONSORT diagram shows participant recruitment, eligibility, enrollment, and selection for inclusion in the study cohort.

**Figure 2.**
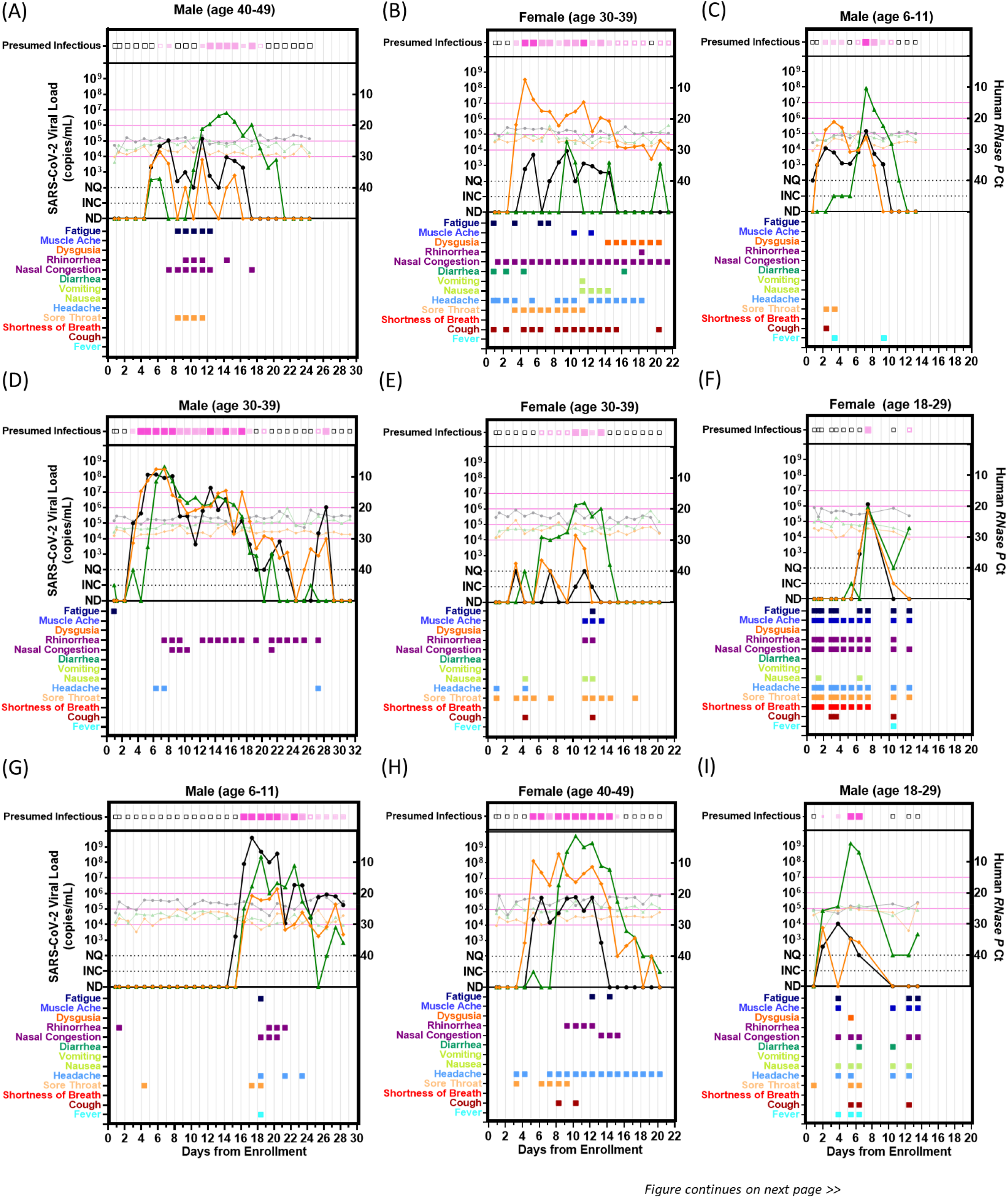
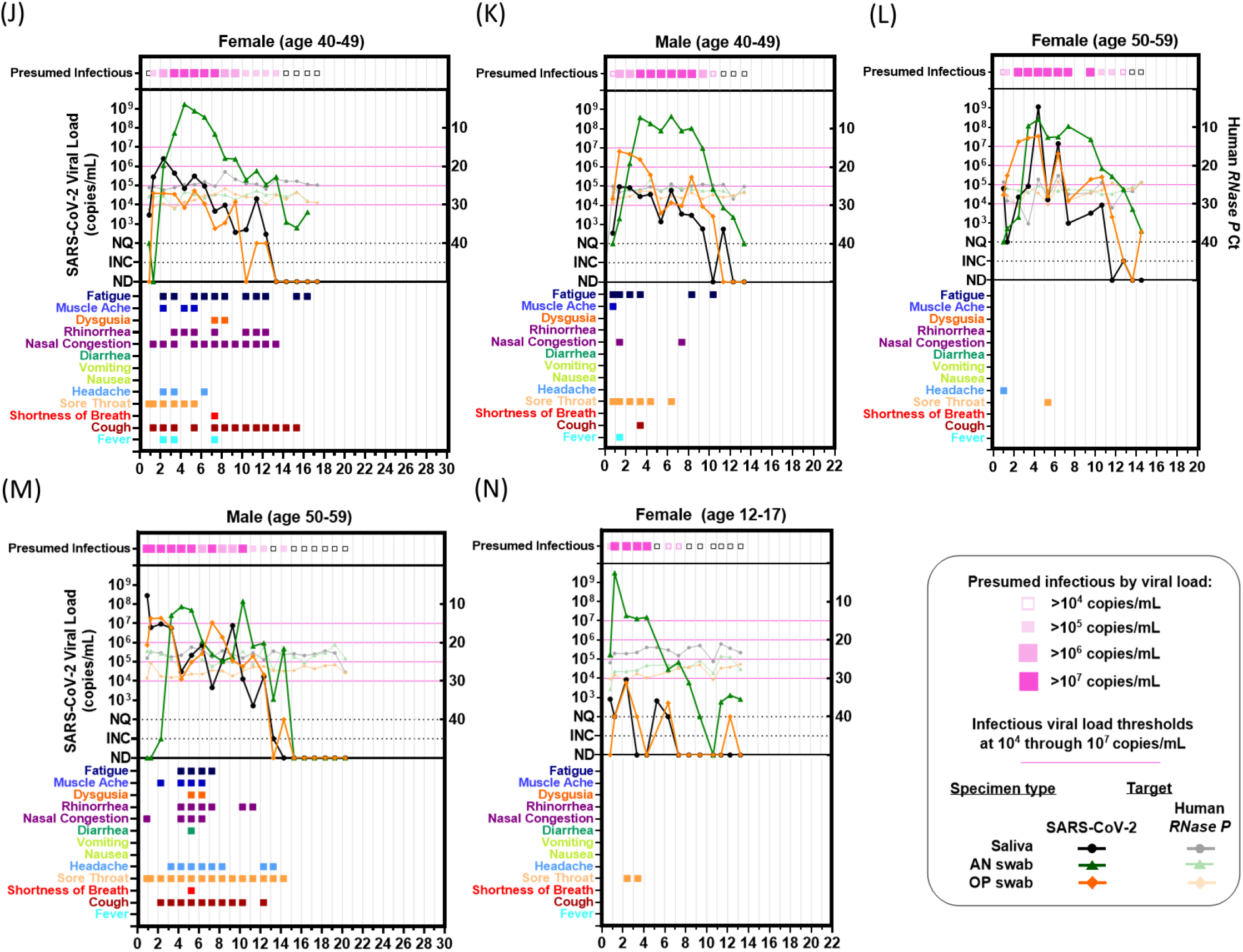
Individual Viral-load Timecourse Measurements from 14 Participants Enrolled at or Before the Incidence of Acute SARS-CoV-2 Infection. Each panel **(A-N)** represents a single participant throughout the course of enrollment. Each panel plots SARS-CoV-2 viral-load measurements left y-axis) and human *RNase P* Ct values (right y-axis). Line colors indicate specimen type: black/grey are saliva, green are anterior-nares (AN) swabs, and orange are oropharyngeal (OP) swabs. Timepoints at which at least one specimen type had presumably infectious viral load (>10^4^-10^7^ copies/mL) are indicated at the top of each plot. Colored boxes below each plot indicate the symptoms reported at each sample-collection timepoint. Each of the 14 participants collected three specimen types throughout the course of acute infection, resulting in 42 viral-load timecourses. Participants collected an average of 15 (±5 S.D.) daily timepoints. ND, not detected; INC, inconclusive result; NQ, virus detected however viral loads below the test LOD (250 copies/mL) and thus not reliably quantifiable for RT-qPCR measurements.

Each day, participants reported symptoms, then self-collected saliva, anterior-nares (hereafter nasal) swab, and posterior oropharyngeal (hereafter throat) swab specimens for RT-qPCR testing in Zymo Research’s SafeCollect devices (CE-marked for EU use), following manufacturer’s instructions(53, 54). Participants collected specimens immediately upon enrollment, then daily every morning upon waking, as morning sample collection has been shown to yield higher viral loads than evening collection(51).

### RT-qPCR Testing for SARS-CoV-2

Extraction and RT-qPCR were performed at Pangea Laboratories (Tustin, CA, USA) using the FDA-authorized *Quick* SARS-CoV-2 RT-qPCR Kit, with results (positive, negative, inconclusive) assigned per manufacturer criteria(55). Additional details can be found in Supplemental Information. This assay has a reported LOD of 250 copies/mL of sample.

### Quantification of Viral Load from RT-qPCR Result

To quantify viral load in RT-qPCR specimens, contrived specimens across a 13-point standard curve (dynamic range from 250 copies/mL to 4.50×10^8^ copies/mL) for each specimen type was generated at Caltech and underwent extraction and RT-qPCR as described above. All three replicates at 250 copies/mL of specimen were detected, independently validating the reported LOD for the assay. For each specimen type, the standard curve generated an equation to convert from SARS-CoV-2 *N* gene Ct values to viral loads in genomic copy equivalents (hereafter copies) per mL of each specimen type. See Supplemental Information for additional details and equations. Positive specimens with viral loads that would be quantified below the assay LOD (250 copies/mL) were considered not quantifiable.

### Viral Sequencing and Lineage/Variant Determination

Viral sequencing of at least one specimen for each participant with incident infection was performed on ANS or OPS specimens with moderate to high viral loads by Zymo Research at Pangea Lab. The sequencing protocol used a variant ID detection workflow that closely resembles the Illumina COVIDSeq NGS Test. See Supplemental Information for details.

### Defining pre-infectious and infectious periods

The “pre-infectious” period is all SARS-CoV-2-positive timepoints prior to the first timepoint in which any specimen type contains viral load greater than the indicated infectious viral load threshold. There are three main methods for defining the infectious period for an individual based on viral loads, all of which will be utilized in this manuscript. First, the infectious period may be defined as the continuous period between the first specimen (of any type) with an infectious viral load until the first timepoint after which no specimen has an infectious viral load(56, 57). Or, to account for viral-load fluctuations, one may instead define an instantaneous infectious period (i.e., an individual is presumed infectious only when at least one specimen type has a viral load above the infectious viral load threshold). Both methods neglect the role of the neutralizing immune response, and the impact of infection stage on viral-culture positivity(31, 58). To account for these factors, the presumed infectious period may be limited to a number of days following symptoms or the first infectious timepoint. In our analyses (**Fig 7**), we performed analyses using all three common definitions. First, we used a “continuous infectious period” whereby a participant is presumed infectious for all timepoints between the first specimen with an infectious viral load and the first timepoint after which no specimens had infectious viral loads. Second, we used an “instantaneous infectious period,” which presumes that a participant is infectious only at timepoints when viral load in at least one specimen type is above the infectious viral load threshold. Third, we presumed that a participant is infectious only for the first 5 days from their first timepoint when at least one specimen type had a viral load above the infectious viral load threshold.

### Statistical Analyses

#### Comparison of Viral-Load Timecourses Across Specimen Types

To quantify the difference between viral-load timecourses, we first aligned each timecourse to the time of collection of the first SARS-CoV-2-positive specimen (of any type) for each participant. Differences between viral loads from the same infection timepoint were quantified (**Fig 3A-B**). We compared both intra- and interparticipant viral-load timecourses: when the lengths of two participant timecourses differed, the longer timecourse was truncated. We then hypothesized that if the viral-load timecourses followed the same time-dependent distribution, then the observed ‘noise’ between these viral-load measurements would be attributable to expected sampling noise.

**Figure 3.**
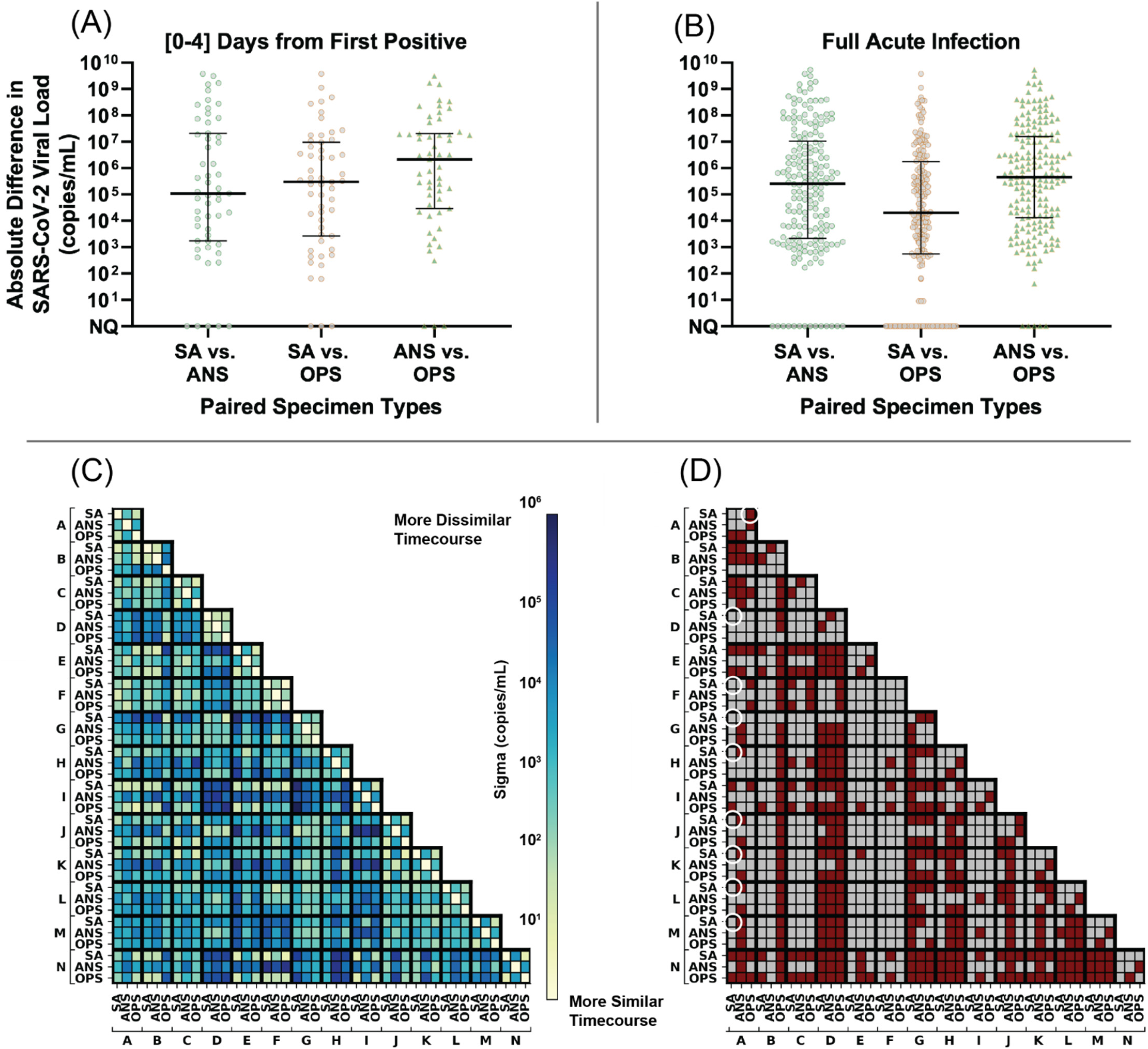
Extreme differences in viral loads across specimen types collected from the same person at the same timepoint for the 14 participants enrolled before or at the incidence of acute SARS-CoV-2 infection. **(A-B)** Absolute differences in viral loads across paired specimen types were calculated as the absolute value of viral load in one specimen type minus another from the same participant at the same specimen-collection timepoint. Black lines indicate median, with interquartile range. Differences are shown for: **(A)** 55 timepoints collected in the first 4 days from the incidence of infection (first positive specimen of any type) in each participant and **(B)** 186 timepoints collected throughout the entirety of acute infection (at least one specimen type from the participant at the timepoint was positive and had quantifiable SARS-CoV-2 viral load; 11 timepoints were positive but not quantifiable). **(C)** Correlation of viral-load timecourses, measured as the standard deviation across paired viral-load timecourses, assuming Gaussian-distributed noise (see Methods “Comparison of Viral-Load Timecourses Across Specimen Types”). **(D)** Statistical significance of the difference in viral-load timecourses between specimens and between participants. Statistically significantly different timecourses are represented as red cells and non-significant comparisons are grey. White circles are called out as examples in the text. Expected sampling noise was estimated by analyzing *RNase P* Ct data from our study **(Fig S4)** and from reference (61). *P*-values were obtained by comparing residuals from observed data and expected sampling noise. Additional method details are shown in **Fig S5**. SA, saliva, ANS, anterior-nares nasal swab, OPS, oropharyngeal swab. Participant labels match **Fig 2** panels (A-N).

Expected sampling noise was estimated as a zero-centered normal distribution fitted on human *RNase P* control target measurements (**Fig S4B**, additional details can be found in Supplemental Information). The distribution of observed ‘noise’ between viral-load measurements was obtained by performing maximum likelihood estimation (MLE) on each pair of viral-load timecourses being compared (**Fig 3C**). We then tested whether observed differences in viral load across pairs of viral-load timecourses could be explained by expected sampling noise alone. *P-*values were then obtained by performing upper-tailed Kolmogorov–Smirnov tests over the differences between the distributions of the observed noise across viral-load timecourses and expected sampling noise. Two-stage Benjamini–Hochberg correction was used to limit the false discovery rate to 5%; viral-load timecourse comparisons with adjusted *P*-values <0.05 were considered statistically significantly different (**Fig 3D**). See Supplemental Information for additional details. Analyses were performed in Python 3.8 using the scipy package(59).

#### Inference of Clinical Sensitivity by Viral-Load Quantification

Inferred clinical sensitivity of a given specimen type and analytical sensitivity was calculated for each timebin of infection as the number of specimens of a given type with viral load above a given LOD divided by all participants considered infected (**Fig 4**) or infectious (**Figs 6-7**) at that timepoint. Confidence intervals were calculated as described in the Clinical Laboratory Standards Institute EP12-A2 User Protocol for Evaluation of Qualitative Test Performance(60). Statistical testing for differences in inferred clinical sensitivity were performed for paired data (comparing performance at two LODs for one specimen type, or at one LOD for two paired specimen types collected by a participant at a timepoint) using the McNemar exact test, and for unpaired data (comparing the performance of one specimen type at one LOD between infection stages) using the Fisher exact test. Analyses were performed in Python v3.8.8.

**Figure 4.**
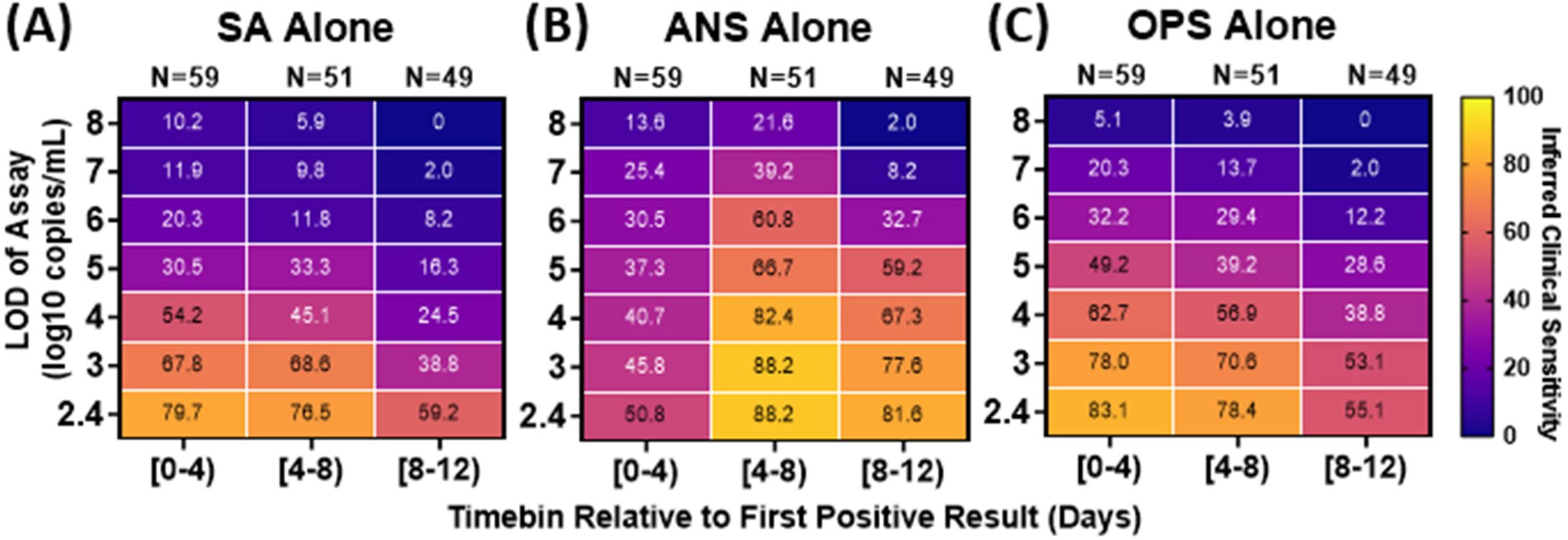
Inferred clinical sensitivity of assays with different LODs to detect infected persons by any single specimen type (A-C) or by computationally-contrived combination specimen types (D-G). Heatmaps show the inferred clinical sensitivity as a function of test limit of detection (LOD) throughout the course of the infection (in 4-day timebins relative to the first positive specimen of any type) for (**A**) SA specimens alone, (**B**) ANS specimens alone, and (**C**) OPS specimens alone. Inferred clinical sensitivity was calculated as the number of specimens of the given type with viral loads greater than the given LOD divided by the total number of specimens collected within that timebin. N indicates the number of timepoints. Only timepoints where at least one specimen type had a quantifiable viral load (≥250 copies/mL) were included. Two-day timebins are shown in **Fig S6**. The performance of combination specimen types using the average (instead of maximum) viral loads is shown in **Fig S7**.

**Figure 5.**
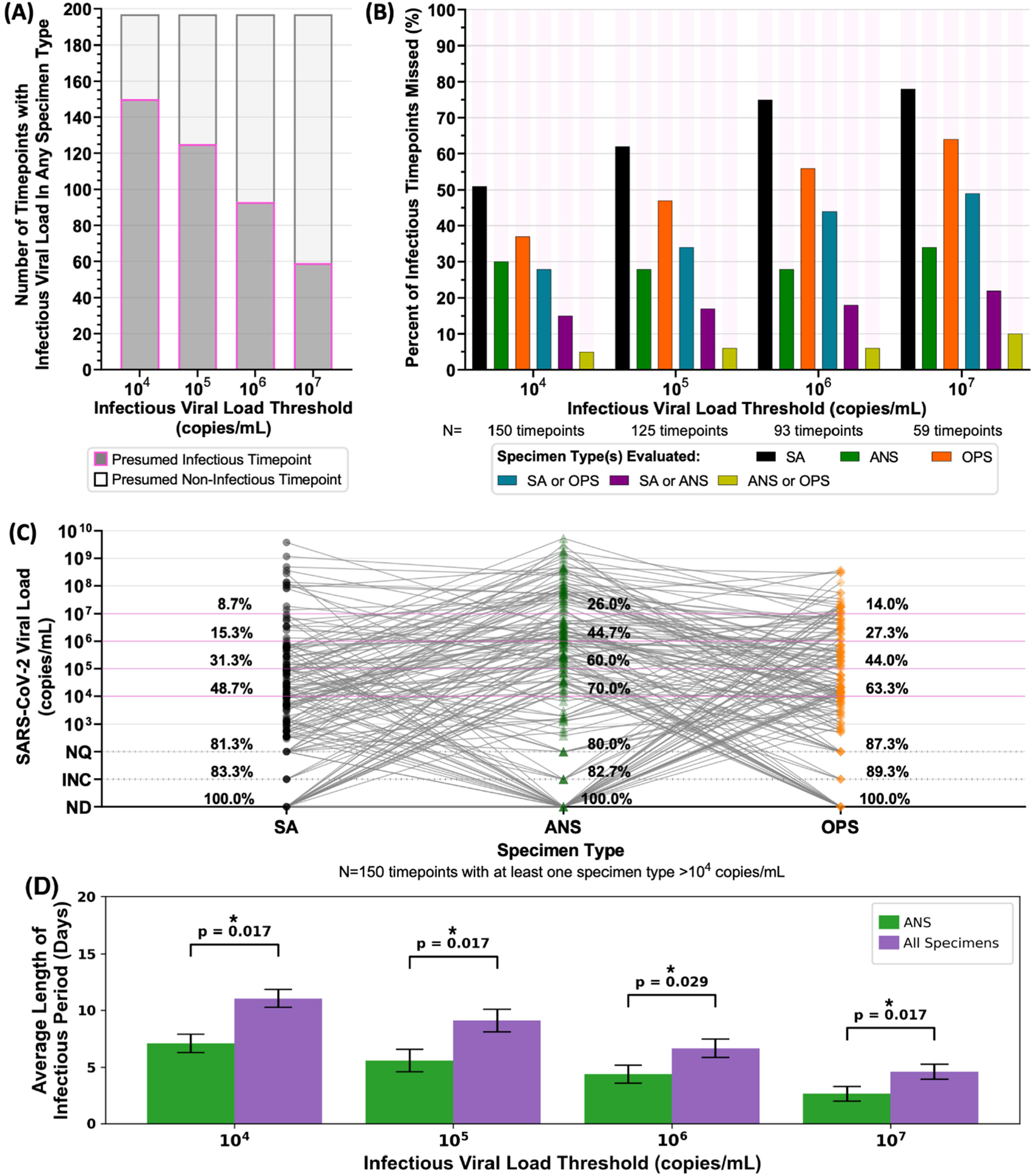
Analyses of Presumed Infectious Viral Loads in Each Specimen Type Using Different Infectious Thresholds. **(A)** Stacked bar plots of the number of timepoints with at least one specimen type above the indicated infectious viral-load threshold (dark grey with magenta outline), and where all paired specimen types collected at a timepoint had viral loads below the infectious viral-load threshold (light grey with black outline). **(B)** Each bar represents the proportion of all infectious timepoints (i.e., saliva or nasal swab or throat swab had a viral load above the infectious viral-load threshold), where the given specimen type or combination of specimen types did not have an infectious viral load. For example, with an infectious viral load threshold of 10^4^ copies/mL, 150 timepoints had an infectious viral load in at least one specimen type: in 105 of those 150 timepoints (70%), the ANS specimen had an infectious viral-load. Therefore, 30% of infectious timepoints would be missed if only the ANS specimen type were evaluated for infectious viral load. Each group of bars provides values for alternate infectious viral-load thresholds, 10^5^, 10^6^, and 10^7^ copies/mL. **(C)** Viral loads of all three specimen types collected by each participant at the same timepoint where at least one specimen type had a viral load above 10^4^ copies/mL (N=150 timepoints). Percentages above each specimen type provide the cumulative proportion of specimens with viral loads at or above each line. Magenta lines indicate possible infectious viral-load thresholds based on literature. (D) Average length of the infectious period when considering only presumably infectious loads in ANS (green) or when considering all specimen types (purple). Error bars are S.E.M. *P*-values were obtained by performing related-sample t-tests for each IVLT. *P*-values were adjusted using two-stage Benjamini–Hochberg correction to account for multiple hypotheses being tested. *ANS, anterior-nares swab; SA, saliva; OPS, oropharyngeal swab; ND, not detected by RT-qPCR; INC, inconclusive result by RT-qPCR; NQ, not quantifiable by RT-qPCR*.

**Figure 6.**
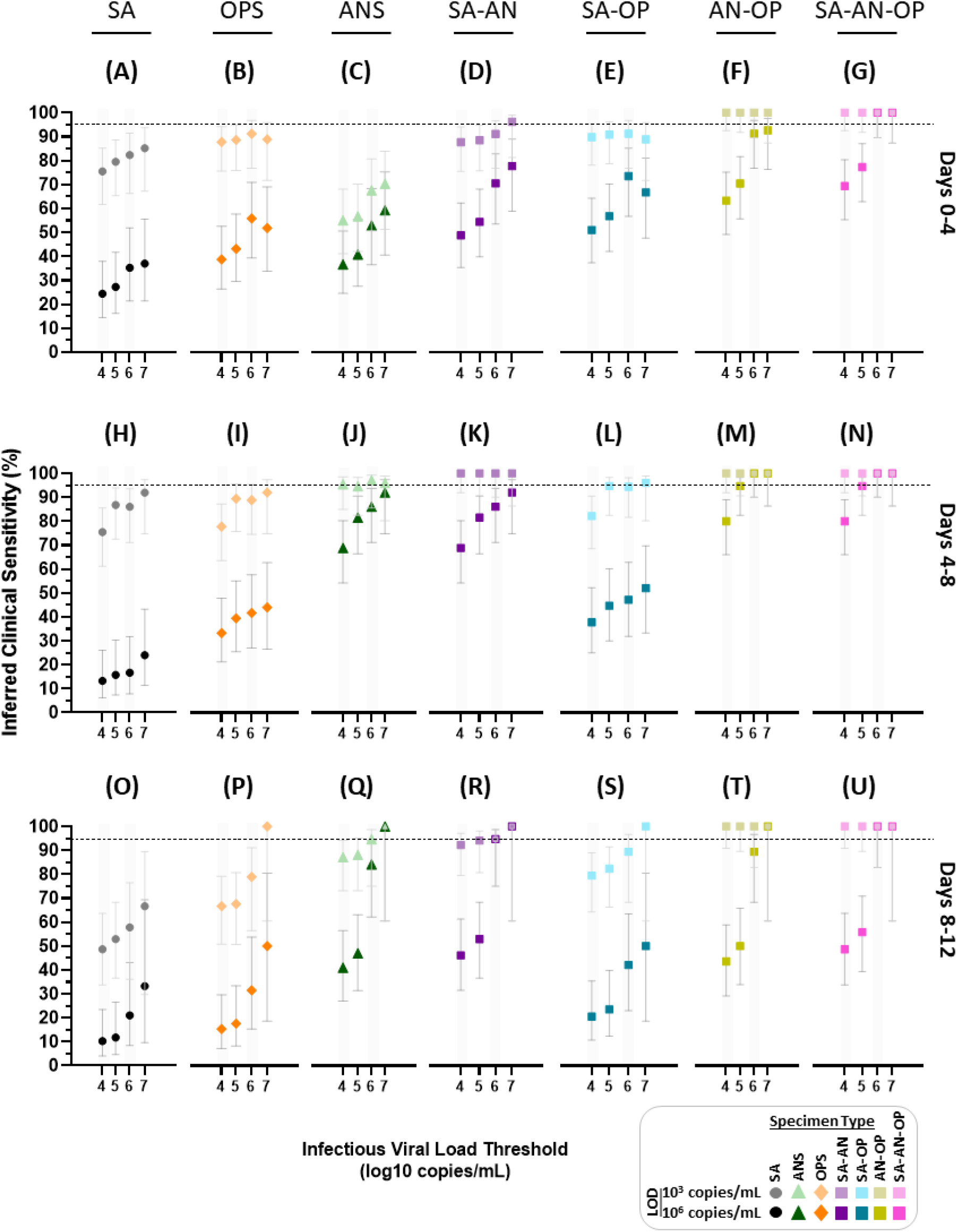
Inferred clinical sensitivity of high- and low-analytical-sensitivity assays to detect presumed infectious individuals by testing single and combination specimen types throughout acute, incident infection. For each 4-day timebin relative to the first SARS-CoV-2 positive specimen (of any type), participants were classified as being presumed infectious if viral load in any specimen type collected at a given timepoint was above an infectious viral load threshold. For a high-analytical-sensitivity assay with an LOD of 10^3^ copies/mL and low-analytical-sensitivity assay with an LOD of 10^6^ copies/mL, the inferred clinical sensitivity was calculated as the number of specimens of that specimen type with a measured viral load at or above the LOD divided by the total specimen-collection timepoints included in that timebin. Error bars indicate the 95% C.I. The viral load of computationally-contrived combination specimen types was taken as the higher viral load of the specimen types included in the combination collected by a participant at a given timepoint. SA, saliva; ANS, anterior-nares swab; OPS, oropharyngeal (throat) swab; SA–AN, saliva-anterior-nares swab combination; SA-OP, saliva– oropharyngeal combination swab; AN–OPS, anterior-nares–oropharyngeal combination swab; SA–AN–OP, saliva-anterior-nares– oropharyngeal combination swab. Inferred clinical sensitivity for LODs from 10^2.4^ to 10^8^ copies/mL shown in **Fig S8**; 2-day timebins are shown in **Fig S9**.

**Figure 7.**
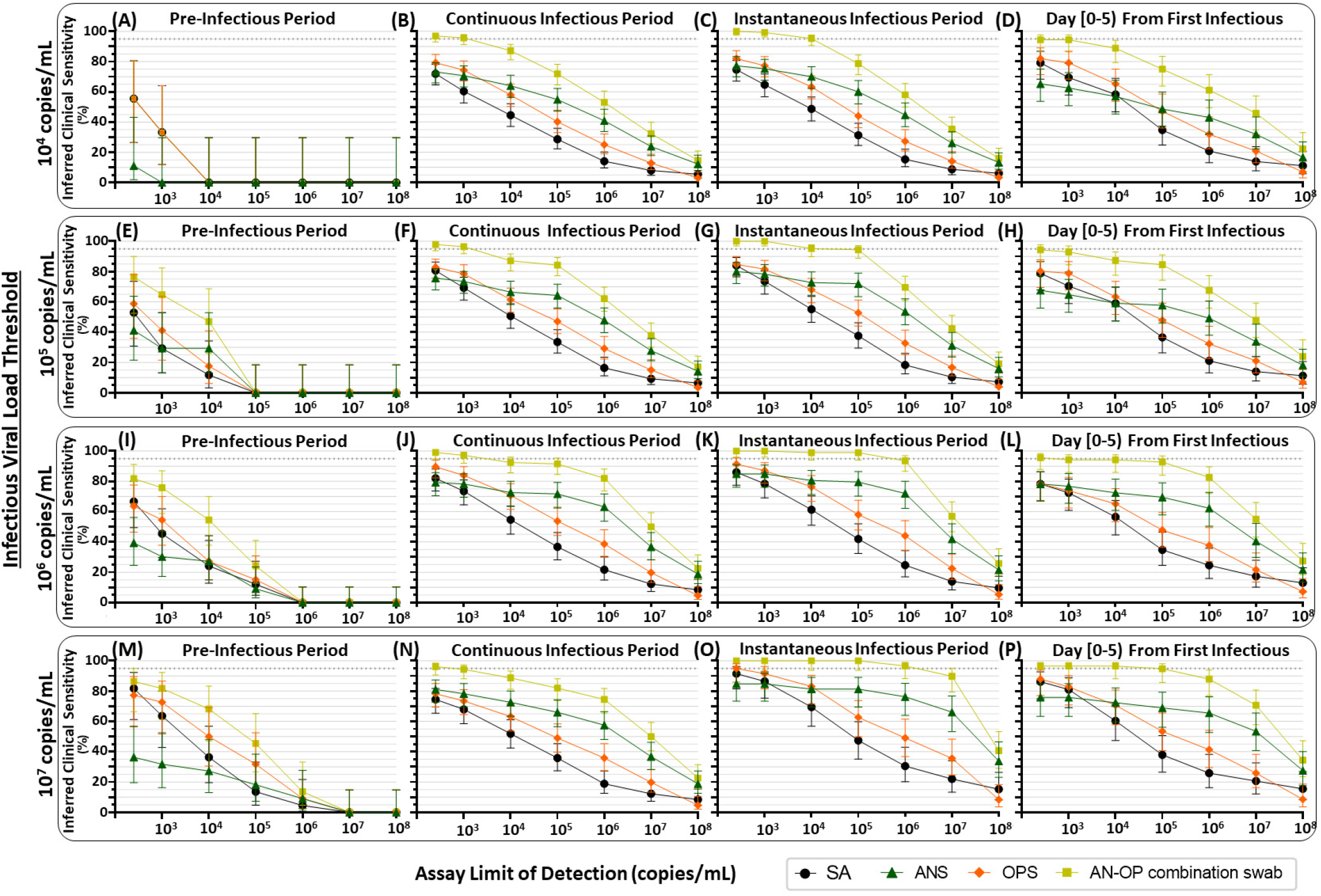
Inferred detection of presumed pre-infectious and infectious individuals at a range of test LODs and with single-specimen tests or AN–OP combination swab specimen type. For each participant, the pre-infectious period was defined as all timepoints with quantifiable SARS-CoV-2 viral load before the first timepoint when at least one specimen type had a viral load above the indicated infectious viral load threshold. We then used three different, common definitions for the infectious period, to assess the robustness of our conclusions. First, we used a “continuous infectious period” whereby a participant is presumed infectious for all timepoints between the first specimen with an infectious viral load and the first timepoint after which no specimens had infectious viral loads. Second, we used an “instantaneous infectious period,” which presumes that a participant is infectious only at timepoints when viral load in at least one specimen type is above the infectious viral load threshold. Third, we presumed that a participant is infectious only for the first 5 days from their first timepoint when at least one specimen type had a viral load above the infectious viral load threshold. These three types of infectious periods were determined for each infectious viral-load threshold: 10^4^, 10^5^, 10^6^, and 10^7^ copies/mL. Each panel provides the inferred clinical performance to detect pre-infectious or infectious individuals, using a given specimen type, for a given assay LOD. Inferred clinical sensitivity was calculated as the number of specimens of each type with a viral load above the assay LOD, divided by the total number of specimens of that type in that period of infection. N indicates the total number of specimens of each type included in the inferred clinical sensitivity calculation. Dotted line indicates 95% inferred clinical sensitivity. *SA, saliva; ANS, anterior-nares swab; OPS, oropharyngeal swab; AN–OP combination swab, predicted combined anterior-nares–oropharyngeal swab specimen type*.

Participants were considered infected from the time of collection of the first SARS-CoV-2-positive specimen (of any type) until negative in all three specimen types by RT-qPCR. Individuals were presumed to be infectious based on whether the viral load in any specimen type were above or below an infectious viral-load threshold (10^4^, 10^5^, 10^6^, or 10^7^ copies/mL) and fulfilled additional definitions described in the Introduction.

Combination specimen types (e.g., anterior-nares–oropharyngeal [AN–OP] combination swab) were computationally-contrived to have either the maximum (**Fig 5-8**) or average (**Fig S7**) viral loads from the specimen types included in the combination that were collected by a participant at that timepoint.

## RESULTS

Among the 228 enrolled participants, incident SARS-CoV-2 infection was observed in 14 participants (**Fig 1**), all of whom were enrolled before or at the start of acute SARS-CoV-2 infection with the Omicron variant of concern. All 14 had received at least one vaccine dose more than 2 weeks prior to enrollment (**Table S1**). From this cohort, 260 saliva, 260 oropharyngeal (throat) swab, and 260 anterior-nares (nasal) swab specimens were collected for viral-load quantification and plotted relative to enrollment in the study (**Fig 2**). All participants additionally took daily rapid antigen tests; an analysis of the antigen testing is in a separate manuscript(52).

Viral-load timecourses in the earliest stage of acute SARS-CoV-2 infection differed substantially among specimen types and participants (**Fig 2**). In only 2 (**Fig 2A, I**) of the 14 participants, viral loads became quantifiable in all three specimen types at the same timepoint; in most (11 out of 14) participants (**Fig 2B,C,D,E,F,G,H,J,K,L,M**), saliva or throat swabs were positive first, while nasal swabs remained negative or at low, inconsistently detectable viral loads for up to the first 6-7 days of infection. However, later in the infection, peak viral loads in nasal swabs were significantly higher than in saliva or throat swabs (**Fig S1**).

Surprisingly, several participants reported zero symptoms on the day of their peak viral loads (**Fig 2A,C,G,N**), all of which were >10^6^ copies/mL. Overall, we found only a weak relationship between viral load and symptoms (**Fig S1-D**). Importantly, individuals had infectious viral loads in 42% of timepoints at which no symptoms were reported (**Fig S1E**).

### SARS-CoV-2 Viral Loads Differ Significantly Between Specimen Types During the Early Period of Infection

We next sought to quantify the magnitude of differences in viral load across paired specimens of different types, to answer three questions: (i) Are differences in viral loads between specimen types large enough to impact detectability by assays with varying analytical sensitivity? (ii) Are differences in viral loads attributable to variability in participant sampling behavior? (iii) Are viral-load timecourses in different specimen types within a person correlated with each other?

First, we calculated the absolute (**Fig 3A**) and relative (fold) differences (**Fig S3**) in viral loads between paired specimens of different types collected by the same participant at the same timepoint. Large (several orders of magnitude) differences in absolute viral loads were observed between paired specimen types for both the first 4 days from the incidence of infection **(Fig 3A)** and all timepoints **(Fig 3B)**. We observed absolute differences of more than 9 orders of magnitude, and all specimen type comparisons had median absolute differences greater than 10^4^ copies/mL, a scale of difference likely to impact the detection of SARS-CoV-2 across different specimen types.

If the observed differences in viral loads between specimen types were the result of variability in sample collection during self-sampling, we would expect the fold differences to be similar to the variability of the human *RNase P* control marker, but *RNase P* Ct measurements were relatively stable for each specimen type collected by participants across their timecourse (**Fig 2, Fig S4**). The average standard deviation in *RNase P* Ct across participants was <1.5 in all specimen types (saliva: 1.37, nasal: 1.42, throat: 1.46) over the entire course of enrollment (**Fig S4B**), which corresponds to, at most, a 2.8-fold change in target abundance. In contrast, most (84%) comparisons between specimen types had greater than a 2.8-fold differences in viral loads (**Fig S2B**), demonstrating the extreme differences in viral load were not due to variability in self-sampling.

Although the differences in viral loads across paired specimens of different types were extreme, we recognized the possibility that the longitudinal timecourse (the rise and fall of viral loads) from different specimen types in a person might still be synchronized. For example, viral loads in one specimen type (e.g., saliva) might be consistently lower than those in another (e.g., ANS), but follow the same pattern throughout acute infection. If this were the case, viral load measured in one specimen type would still be associated with the viral load in another specimen type despite extreme absolute differences.

To test whether timecourses from different specimen types were synchronized, we quantified the correlation between viral-load timecourses for each specimen type collected from a single participant, and across different participants. These intra- and interparticipant correlations are represented as a matrix for the 42 viral-load timecourses (14 participants with three specimen types each, **Fig 3C-D**). The strength of each correlation (**Fig 3C**) was quantified by estimating the standard deviation of pairwise differences in viral load across the two timecourses. The statistical significance of the correlations between viral-load timecourses (**Fig 3D)** was then calculated by comparing the distribution of pairwise differences in viral-load timecourses to a distribution of expected sampling noise.

We found that viral-load timecourses in different specimen types collected by the same individual often do not correlate. In nearly all participants (13 of 14), viral loads in at least two specimen types from the same participant had significantly different timecourses. In 38% of comparisons (16 of 42), we observed significantly different viral-load timecourses for each of the three specimen types from the same individual (**Fig 3D**). In some instances, the viral-load timecourses of specimen types from the same participant were less correlated with each other than with other participants. For example (see white circles in **Fig 3D**), the saliva viral-load timecourse for individual A was not significantly different from the saliva timecourses for participants D, F, G, H, J, K, L, or M, however Individual A’s saliva timecourse was significantly different from the participant’s own throat timecourse.

Within the same individual, throat swab and nasal swab viral-load timecourses were most commonly different (64%, 9 of 14 individuals). Additionally, in 29% (4 of 14) of individuals, saliva and nasal swab viral-load timecourses differed significantly. Finally, despite the proximity of the two oral sampling locations in 21% (3 of 14) of individuals, their own saliva and throat viral-load timecourses were significantly different (**Fig 3D**).

### Clinical Sensitivity to Detect SARS-CoV-2 Infection Strongly Depends on Infection Stage, Specimen Type, and Assay Analytical Sensitivity

Because viral load determines whether an assay with a given analytical sensitivity will reliably yield a positive result, we hypothesized that the extreme differences in viral loads among different specimen types would significantly impact the clinical sensitivity of COVID-19 tests performed on different specimen types during different stages of the infection. To examine the inferred clinical sensitivity to detect SARS-CoV-2 infections as a factor of both specimen type and test LOD, viral-load timecourses were aligned to first detectable viral load and divided into 4-day timebins. We assumed that only viral loads above a given assay’s LOD would reliably yield a positive result. The inferred clinical sensitivity of detecting infected persons by each specimen type and assay LOD during each timebin was calculated as the proportion of specimens with viral loads greater than the assay LOD, divided by all timepoints collected by infected participants in that same timebin (**Fig 4**).

For all specimen types and timebins, testing with a high-analytical-sensitivity assay (LODs of 10^3^ copies/mL) yielded significantly better inferred clinical sensitivity to detect infected persons than testing with a low-analytical-sensitivity assay (LOD of 10^6^ copies/mL) (**Table S4A-I**). During the first 4 days of infection, when individuals are often pre-symptomatic, no single specimen type achieved >90% inferred clinical sensitivity with any LOD (**Fig 4A-C**), suggesting that no single specimen type will reliably provide early detection of infection with the Omicron variant.

In the first 4 days, nasal swabs generally had the poorest inferred clinical sensitivity of all three specimen types. Even with a high-analytical-sensitivity assay (LOD of 10^3^ copies/mL), nasal swabs were predicted to miss more than half (54%) of timepoints from infected persons. Saliva and throat-swab specimens had significantly better inferred clinical sensitivity than nasal swabs when a high-analytical-sensitivity assay (LOD of 10^3^ copies/mL) was used, and worse (but not significantly) when a low-analytical-sensitivity assay (LOD of 10^6^ copies/mL) was used (**Table S4J-Z)**.

As infection progresses to days 4-8, individuals are more likely to become symptomatic. Inferred nasal-swab performance improved significantly during days 4-8 (**Table S4 AH-AN**) and became significantly better than saliva and throat swabs at LODs of 10^3^ copies/mL and above (**Table S4AO-BB**). This improvement can be attributed to the rise to high viral loads in nasal swabs during this period: SARS-CoV-2 was not detected in almost half of nasal swabs in days 0-4, but in days 4-8, more than half of nasal swabs had high viral loads (>10^6^ copies/mL) (**Fig S1I,J**). In contrast, during both timebins, more than half of all saliva or throat-swab specimens had viral loads below 10^6^ copies/mL, and thus detection using saliva or throat swabs was more dependent on assay LOD (**Fig S1I,J**).

### Differences in Viral Loads Among Specimen Types Hinders Detection of Presumably Infectious Individuals When Tests Utilize Single Specimen Types

Prompt identification of individuals who are or will become infectious can prevent further transmission. We next compared the ability of each specimen type and assay analytical sensitivity to detect presumably infectious individuals. An individual was presumed to be infectious if the viral load in any specimen type collected from that participant at a given timepoint was above an infectious viral load threshold. We performed separate analyses for four well-accepted infectious viral-load thresholds (log values of 10^4^ to 10^7^ copies/mL) to test the robustness of our conclusions.

We found that because of the extreme differences in viral-load timecourses, a presumed non-infectious viral load in one specimen type did not reliably indicate that a participant would have presumed non-infectious viral loads in all specimen types. At the highest infectious viral-load threshold (10^7^ copies/mL), a presumed non-infectious viral load in one specimen type (**Fig 5A**) correctly inferred the participant did not have an infectious viral load in any specimen type collected at that timepoint 70% of the time (138 of 197 timepoints). In contrast, at the lowest infectious viral-load threshold (10^4^ copies/mL), a presumed non-infectious viral load in one specimen type correctly inferred a non-infectious participant only about 24% of the time (47 of 197 timepoints).

Across infectious viral-load thresholds, we saw a pattern that suggested combination specimen types might capture more presumably infectious timepoints than single specimen types (**Fig 5B-C**), as 90-95% of timepoints with a presumed infectious viral load in any specimen type had infectious viral loads in either nasal swab or throat swab. This complementarity suggested that a nasal–throat combination swab specimen type could be superior for detecting nearly all infectious individuals.

We interrogated this complementarity between nasal and throat swabs by comparing the viral loads of the three specimen types at each of the 150 timepoints in which at least one specimen had viral loads above a 10^4^ copies/mL infectious viral-load threshold (**Fig 5C**). We found that 52% of individuals with presumed non-infectious viral loads in saliva, 38% of individuals with presumed non-infectious viral loads in throat swabs, and 30% of individuals with presumed non-infectious viral loads in nasal swabs actually had presumably infectious viral loads in another specimen type at the same timepoint. In some cases, high-analytical-sensitivity testing could capture individuals with infectious viral loads in specimen types other than the one tested. However, 19% of saliva, 20% of nasal swab, and 13% of throat swab specimens had either undetectable or unquantifiable viral loads while another specimen type in the same individual had an infectious viral load (**Fig 5C**). In such cases, testing a single specimen type even with a very-high-analytical-sensitivity assay (e.g., LOD of 250 copies/mL) would not reliably detect a presumably infectious person.

Given that the infectious periods for different specimen types were often asynchronous, considering infectiousness in all three specimen types yielded a significantly longer infectious period than if only ANS viral loads were considered (**Fig 5D**) across all infectious viral load thresholds. We also found that the infectious period in ANS and OPS together was longer than any other combination of two specimen types, and similar to that of all three specimen types. These results suggest that testing only single specimen types (such as ANS) may fail to detect individuals with infectious viral loads in untested specimen types.

### Inferring Detection of Infectious Individuals by Specimen Type and Assay Analytical Sensitivity Across Infectious Viral-Load Thresholds

Having observed that a person can have low viral loads in one specimen type while having high and infectious loads in another type prompted us to question how well each specimen type and assay LOD would impact the detection of infectious individuals at different stages of the infection. We binned timepoints into 4-day bins and assessed the ability of each specimen type to detect presumably infectious individuals using assays with either high-(LOD 10^3^ copies/mL) or low-(LOD 10^6^ copies/mL) analytical sensitivity in each bin (**Fig 6**).

Regardless of specimen type, the inferred clinical sensitivity of both high and low-analytical-sensitivity assays to detect presumed infectious individuals typically increased as the infectious viral-load threshold increased. Improved clinical sensitivity at higher infectious viral-load thresholds was most pronounced for assays with LODs of ≥10^6^ copies/mL. This pattern is intuitive; specimens with viral loads above the infectious viral-load threshold but below the LOD are presumed infectious but missed by the assay, resulting in poor inferred clinical sensitivity. Increasing the infectious viral-load threshold would exclude those specimens from being presumed infectious, thereby resulting in better inferred clinical sensitivity (**Fig 5**).

Three major patterns in the specimen types were consistent regardless of the infectious viral-load threshold, so for simplicity the rest of this section describes inferred clinical performances and statistical comparisons using only an infectious viral-load threshold of 10^5^ copies/mL. First, even when tested with a high-analytical-sensitivity assay, no single specimen type achieved >95% inferred clinical sensitivity to detect presumed infectious individuals (**Fig 6A-C**). Second, because the rise in nasal-swab viral load was delayed relative to saliva or throat swab in most participants (**Fig 2**), nasal swabs had significantly worse performance than saliva and throat swabs during days 0-4 (**Table S4BS-BU**). At an assay LOD of 10^3^ copies/mL, the inferred clinical sensitivity of nasal swabs was only 57% (**Fig 6C**). This suggests that nasal-swab testing, even with high analytical sensitivity, would miss approximately 43% of presumed infectious individuals the first 4 days of infection. Third, from days 4-8 of infection, when nasal-swab viral loads increased rapidly in many participants (**Fig 2, Fig 4**), nasal swabs had significantly higher inferred clinical sensitivity regardless of LOD (**Fig 6C,H-J; Table S4BU,BV)** across LODs **(Table S4BW-BZ)**.

### Combination specimen types are inferred to significantly improve the clinical sensitivity to detect infected and infectious individuals with assays of any analytical sensitivity

The extreme differences and lack of correlation in viral loads among specimen types as well as the poor performance of all three specimen types in all timebins and all test LODs led us to hypothesize that combination specimen types might achieve better clinical sensitivity. We generated computationally-contrived specimen types representing combinations of specimen types. For each timepoint, the viral load of a combination specimen type was the highest viral load of any single specimen type included in the combination. We then inferred the clinical sensitivity of these combination specimen types to detect infectious individuals with assays of different analytical sensitivities for each timebin (**Fig 4D-G**). The high clinical sensitivity of throat swabs days 0-4, and of nasal swabs at days 4-8 suggested complementarity. Complementarity was further supported by nasal and throat swabs having the most extreme differences in viral load (**Fig 3A,B**), that that many individuals had significantly different nasal-swab and throat-swab viral-load timecourses (**Fig 3D**). Moreover, rarely did individuals have infectious viral loads in saliva alone (**Fig 5, Table S2**).

Indeed, the nasal–throat combination swab had higher clinical sensitivity to detect infected individuals than any single specimen type, at most LODs (**Fig S6**). This nasal-throat combination specimen type (**Fig 6F**) was also inferred to perform significantly better than all single specimen types (**Fig 6A-C**) at detecting presumed infectious individuals during the first 4 days of infection, and significantly better than saliva (**Fig 6H,O**) and throat swab (**Fig 6I,P**) during later stages of infection (**Fig 6M,T, Table S4CA-CJ**). In addition, the nasal–throat combination swab had significantly better inferred performance than nasal swabs when tested with a low-analytical-sensitivity assay during days 4-8 (**Fig 6J,M**). The combination of all three specimen types (**Fig 6G,N,U**) would by definition capture all presumed infectious individuals. However, this combination type never had a significantly higher inferred clinical sensitivity than nasal–throat combination swab (**Table S4CM-CR**).

### Performance of Specimen Types and Analytical Sensitivities in the Pre-infectious and Infectious Periods

For public-health purposes, understanding assay performance during the pre-infectious and infectious periods, rather than in timebins relative to the rarely-captured incidence of infection, is more informative and actionable. Therefore, we next evaluated the performance of each single specimen type and the nasal–throat combination swab for each assay LOD during the presumed pre-infectious and infectious periods (**Fig 7**). To ensure our conclusions were robust we compared the results of our analysis across three definitions of the infectious period: a “continuous” infectious period, an “instantaneous” infectious period), and a “day [0-5]” infectious period (only the first 5 days after an initial presumed infectious specimen (see Methods).

At all infectious viral-load thresholds above 10^5^ copies/mL, the nasal–throat combination swab had the highest inferred clinical sensitivity of any specimen type to detect pre-infectious individuals (**Fig 7E,I,M**). In all cases where the assay LOD was at least 2 orders of magnitude lower than the infectious viral-load threshold, there were more than 10 detectable specimens available for comparison of inferred clinical sensitivity and nasal–throat combination swab was inferred to perform significantly better than nasal swab alone (**Table S4CS-DT**). With an infectious viral-load threshold of 10^4^ copies/mL, fewer pre-infectious timepoints were available for analysis. In this case, we see that nasal swabs had very low performance, but no specimen type emerged as optimal (**Fig 7A**).

Three additional trends held across all infectious viral-load thresholds and all definitions of the infectious period. First, nasal swabs had similar performance to saliva and throat swabs when testing with high-analytical-sensitivity assays (LODs at or below 10^3^ copies/mL), except when infectious period is defined as the 5 days following the first infectious specimen. This definition selects earlier timepoints, prior to the rise in nasal swab viral loads (**Fig 2, Fig 5C**) so nasal-swab testing had lower inferred clinical sensitivity to detect both infected (**Fig 4B**) and infectious (**Fig 6C**) individuals. Second, as noted previously (**Fig 4**), nasal-swab performance for the detection of infectious individuals was more robust to differences in assay LOD than saliva and throat swabs because nasal-swab loads tended to be either very low or very high (>10^6^ copies/mL), whereas saliva and throat swabs tended to fluctuate between 10^4^ to 10^7^ copies/mL (**Fig S1D,E**). Furthermore, in all but one comparison (**Fig 7D**) nasal swabs were inferred to have higher performance than saliva or throat swab alone when tested with lower analytical sensitivity assays (LODs at and above 10^5^ copies/mL). Third, a nasal–throat combination swab always had the highest inferred clinical sensitivity at all LODs.

## DISCUSSION

In 14 individuals enrolled before or at the incidence of acute infection, we observed extreme and statistically significant differences in SARS-CoV-2 viral loads among three common respiratory specimen types (saliva, anterior-nares [nasal] swab, and oropharyngeal [throat] swab) collected at the same timepoint from the same individual. In all 14 individuals we also observed that the viral-load measurements in different specimen types followed significantly different longitudinal timecourses. These intra-participant differences were as extreme as those observed between participants (**Fig 3C-D**). The differences in viral load resulted in significantly different inferred clinical sensitivities to detect both infected and infectious individuals depending on the infection stage, specimen type, and analytical sensitivity (limit of detection) of the assay. We conclude that unlike infections where a single specimen type is typically sampled to test for virus (e.g., HIV in blood), SARS-CoV-2 viral load only describes the state of the specimen type tested, not the general state of the individual’s infection.

A person can have high and presumably infectious viral loads in one specimen type but low or even undetectable loads in another specimen type at the same time point. Thus, defining infectiousness based on assessment of only one specimen type(31, 32, 35, 37, 38, 62-68) likely underestimates the full infectious period, particularly if nasal swabs (which typically exhibit infectious viral loads days after oral specimen types) alone are used. Relatedly, policies guiding isolation time that are based on estimates of the infectious period from a single specimen type may result in premature release of infectious individuals from isolation. Our results also suggest that field evaluations of diagnostics to detect infectious individuals that use a single specimen type as the comparator assay(64, 69-75) are likely to overestimate the clinical sensitivity of the test being evaluated.

Because of the extreme differences in viral-load patterns in the early and pre-infectious periods of infection, of the three specimen types considered here, none is optimal for detecting Omicron. However, nasal swab was the poorest specimen type for detection in the first 4 days of infection. In most participants, we observed a delay in nasal-swab viral loads relative to oral specimens similar to what has been observed previously(13, 21, 76) with earlier SARS-CoV-2 variants. In our study, 12 of 14 participants (86%) were either negative in nasal-swab specimens or had nasal-swab viral loads below 250 cp/mL at the incidence of infection (the first day viral RNA was detected in any specimen type). In 3 of these 12 participants (25%), nasal-swab viral loads were either undetectable or inconclusive for more than 5 days (**Fig 1B,C,H**). Because of the delay in nasal-swab viral loads in the first days of infection, the inferred clinical sensitivity of nasal swabs at the beginning of infection was low (<60%), even with high-analytical-sensitivity assays. This delay in nasal-swab viral load and the resulting poor clinical sensitivity of nasal-swabs raises concerns about the performance of diagnostic tests that use nasal specimens and also any diagnostic assays that have been validated against tests that use nasal specimens.

Furthermore, we found that low-analytical-sensitivity testing was inferred to have poor performance for early detection of infected individuals, regardless of the specimen type used. High-analytical-sensitivity assays (e.g., LODs of ≤10^3^ copies/mL) were inferred to improve clinical sensitivity in all specimen types and at all stages of infection. We also found that even with high-analytical-sensitivity testing, none of the three specimen types considered here were optimal for detection of presumed infectious individuals (based on viral-load thresholds of 10^4^ to 10^7^ copies/mL or greater in any specimen type). Of the three single specimen types, nasal-swab testing was inferred to miss the lowest proportion of presumed infectious individuals overall; yet nasal swabs still missed at least a quarter of all presumably infectious timepoints because of high viral loads in oral specimen types (**Fig 5-7**). The failure to detect presumed infectious individuals was inferred to be even worse when using tests of low analytical sensitivity. To assess this point directly, rapid antigen testing results for a broader cohort from this study population are reported in a separate paper(52).

Testing with combination specimen types (e.g., sampling from both the throat and anterior nares) was inferred to yield significantly improved clinical sensitivity to detect both infected (**Fig S6, S7**) and presumed infectious individuals (**Fig 6-7**) than any single specimen type, regardless of whether the combination specimen type was assumed to have the maximum or the average viral load of constituent specimen types (**Fig S7**). Combination swabs have high acceptability(77), and are already common in many regions of the world. In the U.K., the National Health Service website even states that PCR tests that rely only on nasal swabbing will be “less accurate” than those with a combined nose and tonsil swab(78, 79). The U.K. also uses a combination throat–nasal swab for rapid antigen testing. However, despite hundreds of emergency use authorizations (EUAs) that the U.S. FDA has issued for diagnostics that detect SARS-CoV-2(80), including 280 molecular tests and 51 antigen rapid diagnostic tests, none use a combination specimen type.

Our results explain why studies comparing single and oral-nasal combination specimen types have generally shown that combination specimens are either equivalent(25, 81-85) or superior(86-91) to single specimens. Importantly, in nearly all studies evaluating the use of combination swabs, or evaluating combination swab antigen rapid diagnostic tests using a combination swab RT-PCR as reference(32, 50), sample collection began *after* the onset of COVID-like symptoms and/or *after* an initial positive test (usually by nasal swab); thus, they likely did not sample the earliest days of infection, which is the period when we found the greatest benefit of sampling with saliva or a throat swab. One prospective cohort study that did begin testing early (using presymptomatic and asymptomatic close contacts) and used combination oropharyngeal-nasal swabs with an RT-qPCR assay as reference to evaluate two antigen rapid diagnostic tests(40) found a similar clinical sensitivity to detect presumed infectious individuals (∼85-90%) with this combination swab specimen type as what we inferred for a combination swab specimen type based on the viral loads in each specimen type individually tested with a moderate- or low-analytical-sensitivity assay.

We note three main limitations. First, although this is the most comprehensive study of complete viral loads in multiple specimen types to date, data are from a limited number of individuals and demographic. Obtaining early viral-load timecourses from these 14 individuals required enrollment and daily testing of 228 participants for a total of 6,825 RT-qPCR tests. Future studies for new SARS-CoV-2 variants and new respiratory viruses should ideally involve multi-institution partnerships to enroll a diverse cohort from a broad geographic range. Second, we presumed infectiousness based on viral-load thresholds in three specimen types; we did not perform viral culture on these specimens (and acknowledge that specimen types not collected here could have contained infectious viral loads(92)). Third, Omicron remains a relevant variant more than a year after its emergence, but additional variants will continue to develop and may exhibit different patterns in their viral-load timecourses by specimen type, so similar community-based studies will be needed to identify optimal testing methods for new variants (and emerging respiratory viruses).

Viral loads are used in many clinical and basic-science contexts, including diagnostics, epidemiological models, clinical trials, and studies of human immune response. Our results show that early in SARS-CoV-2 infection, viral load cannot be defined for a person, only for a specific specimen type within a person. Thus, when viral load studies or viral detection studies are performed with only single specimen type, the results should be interpreted while considering the heterogeneity of viral loads across specimen types. Additional quantitative longitudinal studies of differences in viral loads in multiple specimen types starting immediately at the incidence of infection are needed for new emerging variants and new respiratory viruses. In the absence of such studies, combination specimen types and tests with high analytical sensitivity are likely to be the most robust approaches for earliest detection and for the design of studies seeking to assess infection status or presence of infectious virus.

## Data Availability

The data underlying the results presented in the study can be accessed at CaltechDATA: https://data.caltech.edu/records/20223.

## Acknowledgments

This work was funded in part by a grant from the Ronald and Maxine Linde Center for New Initiatives at the California Institute of Technology (to RFI), a grant from the Jacobs Institute for Molecular Engineering for Medicine at the California Institute of Technology (to RFI), and a DGSOM Geffen Fellowship at the University of California, Los Angeles (to AVW). We sincerely thank the study participants for making this work possible. We thank Lauriane Quenee, Grace Fisher-Adams, Junie Hildebrandt, Megan Hayashi, RuthAnne Bevier, Chantal D’Apuzzo, Ralph Adolphs, Victor Rivera, Steve Chapman, Gary Waters, Leonard Edwards, Gaylene Ursua, Cynthia Ramos, and Shannon Yamashita for their assistance and advice on study implementation and/or administration. We thank Jessica Leong, Ojas Pradhan, Si Hyung Jin, Emily Savela, Bridget Yang, Ekta Patel, Hsiuchen Chen, Paresh Samantaray, Zeynep Turan, Cindy Kim, Trinity Lee, Vanessa Mechan, Katherine Stiefel, Rosie Zedan, Rahulijeet Chadha, Minkyo Lee, and Jenny Ji for volunteering their time to help with this study. We thank Prabhu Gounder, Tony Chang, Jennifer Howes, and Nari Shin for their support with recruitment. Finally, we thank all the case investigators and contact tracers at the Pasadena Public Health Department and Caltech Student Wellness Services for their efforts in study recruitment and their work in the pandemic response.

## Supplemental Information for

### Supplemental Materials and Methods

#### Study Participants

All adult participants provided written informed consent, and minors provided assent and their legal guardian provided written permission. Individuals were eligible for enrollment if someone in their home had recently (within 5 days) become positive for SARS-CoV-2, or if they had a recent known exposure to a person suspected to be SARS-CoV-2-positive. All participants had to be 6 years of age or older and fluent in English.

#### Extraction and RT-qPCR

Participants packaged their specimens each morning for transport by medical courier to Pangea Laboratories in Tustin, CA, USA. Most specimens were received at the facility within 10 hours of collection; some specimens were received at the facility ∼24-48 hours after donation due to transport delays. Most specimens were extracted and run in RT-qPCR within a few hours of arrival to the facility. Extraction and RT-qPCR operators and supervisors (at Pangea Laboratory) were blinded to which participant a specimen originated from, as well as the infection status and test results of participants.

Extraction and RT-qPCR were performed using the FDA-authorized Quick SARS-CoV-2 RT-qPCR Kit.^58^ which extracts nucleic acids using the Quick-DNA/RNA Viral MagBead Kit (Zymo Research, Catalog #R2141) followed by amplification of three target regions within the SARS-CoV-2 *N* gene.

A specimen was considered inconclusive if the human *RNase P* Ct value was >40 or not detected. If *RNase P* had a Ct < 40, then for a SARS-CoV-2 *N* gene target Ct value <40 the sample was considered positive. If the SARS-CoV-2 target Ct value was 40-45 it was considered inconclusive, and if >45 or not detected it was considered negative.

#### Quantification of Viral Load from RT-qPCR Result

To quantify viral load in RT-qPCR specimens, a 9-point standard curve was generated at Caltech using dilutions from a commercial heat-inactivated SARS-CoV-2 particles (BEI Cat. N4-52286 Lot 70034991). To achieve higher concentrations and greater dynamic range in the standard curve, volume from a participant saliva specimen previously quantified to have a viral load of 6.44×10^9^ copies/mL^53^ was used to generate 4 additional points. Diluted particles or volume from the participant specimen was spiked into pooled matrix from freshly collected SA, ANS, or OPS specimens from SARS-CoV-2 negative donors, collected as described above. Specimens were then shipped to Pangea Laboratories (concentrations blinded) for extraction and RT-qPCR testing. Three of three replicates at 250 copies/mL of specimen were detected, independently validating the reported LOD for the assay.

From the dynamic range of the standard curve (250 copies/mL to 4.50×10^8^ copies/mL), the following equations were used to convert RT-qPCR SARS-CoV-2 N gene Ct value to viral load in genomic copy equivalents (copies) per mL of each specimen type:

- Viral Load in copies/mL saliva = 2^(**Ct** - 42.374)/-0.8973^
- Viral Load in copies/mL buffer for nasal swabs = 2^(**Ct** - 43.050)/-0.9282^
- Viral Load in copies/mL buffer for oropharyngeal swabs = 2^(**Ct** - 43.903)/-0.9653^

Positive specimens with viral loads that would be quantified below the assay LOD (250 copies/mL) were considered not quantifiable, as amplification and resulting Ct values become noisy at these very low viral loads.

#### Viral Sequencing and Lineage/Variant Determination

Whenever possible, we sequenced the putative index case’s highest viral load nasal-swab specimens. When this was not possible (e.g., if the index case was not enrolled, or the index case’s highest viral load nasal-swab specimen was insufficient for sequencing, or limitations in available specimen volume), we chose an alternate high viral load (viral load <2×10^4^ copies/mL) nasal or oropharyngeal swab specimen from the index case or a secondary case in the household.

All sequencing was performed by Zymo Research at Pangea Lab using a variant ID detection workflow that closely resembles the Illumina COVDISeq™ NGS Test (EUA).^59,60^ In brief, RNA extracted from samples underwent cDNA synthesis using random hexamers according to the manufacturer’s recommendation (Illumina, Catalog #20043675).

The SARS-CoV-2 virus genome was amplified using primers designed to tile across the full sequence length as originally described by the ARTICnetwork (https://artic.network/ncov-2019). Amplicons containing the SARS-CoV-2 viral genome fragments were then pooled and subjected to tagmentation to further fragment and tag amplicons with adapter sequences. Adapter-tagged amplicons then underwent a second round of PCR amplification using a PCR master mix and unique index adapters. The indexed libraries were then pooled and cleaned up for downstream sequencing.

Finished libraries were sequenced on an Illumina MiniSeq using a PE 100 bp read configuration to a depth of approximately 100,000 reads per library. Illumina sequence reads were converted from bcl to fastq files, adaptor trimmed, then quality filtered using standard parameters. Variant calls as described by Phylogenetic Assignment of Named Global outbreak LINeages software 2.3.2 (github.com/cov-lineages/pangolin) were made using a custom bioinformatics data analysis pipeline developed by Zymo Research.

#### Shuffled Viral-Load Timecourses and Data Validations

In addition to controls built into the study design (e.g. specimen have barcodes specific to each specimen type, barcodes are confirmed to be the expected specimen type when packaging specimen-collection materials prior to delivery to participants, participants take and package specimen types in a specific order during each timepoint, and the receiving laboratory assessed arriving specimen for the presence of a swab), we assessed mathematically whether the observed viral loads were likely to come from viral-load timecourses of their designated specimen type, or whether they could have been switched between specimen types. We assessed the correlation between the viral load for a given specimen at a timepoint and either the viral load in the same specimen type or the viral load from a different, randomly selected specimen type at the following timepoint (**Fig S3**), for all measurements. The correlation between viral-load measurements from randomly selected specimen types is significantly different (*P*<0.001) from the correlations between viral-load measurements from the same specimen type (**Fig S3C**). Erroneously assigned specimen types would yield similar (*P*>0.01) correlations for both randomized and non-randomized viral-load timecourses. The analysis showed greater standard deviation for shuffled compared with unshuffled viral-load timecourses, suggesting that all specimens were correctly assigned to specimen type by participants.

#### Estimations of Sample Noise with RNase P

To estimate expected sampling noise that would affect viral-load measurements in each specimen type, we examined RT-qPCR Ct measurements of the human *RNase P* control target in the same specimen type from each of the 14 participants in this cohort (**Fig 2**; **Fig S4B**). The standard deviation of the *RNase P* Ct was calculated for each timecourse and then averaged over all 14 participants: the average standard deviation of *RNase P* Ct for saliva specimens was 1.37, nasal-swab specimens was 1.42, and oropharyngeal swab specimens was 1.46 (**Fig S4B**). We then used the average standard deviation of *RNase P* Ct across all three specimen types (1.42 Ct) as the overall estimate of sampling noise in all viral-load measurements, which is consistent with the standard deviation (1.7 Ct) of SARS-CoV-2 *N2* gene Ct values in two MT nasal-swab specimens collected immediately in sequence in a separate study.^66^

#### Alternate Viral Load Calculation for Computationally Contrived Combination Specimens

We recognize that specimen-collection and processing factors (e.g., buffer volumes, type and carrying capacity of swabs), may cause dilution effects that would impact the viral load for combination specimen types. To account for this, we also performed an analysis where the viral load of a computationally-contrived combination specimen was calculated as the average (rather than maximum) viral load of paired single specimen types in each combination (**Fig S7**). Using the average introduced at most a 2- or 3-fold correction for the two- or three-specimen combinations, respectively, because viral loads differed by orders of magnitude (**Fig 3**). Clinical sensitivities of combination specimen types remained similar (**Fig S7I-J**) to those calculated in **Fig 4** and the nasal–throat combination swab remained superior with this alternate calculation (**Fig S7F**).

**Figure S1.**
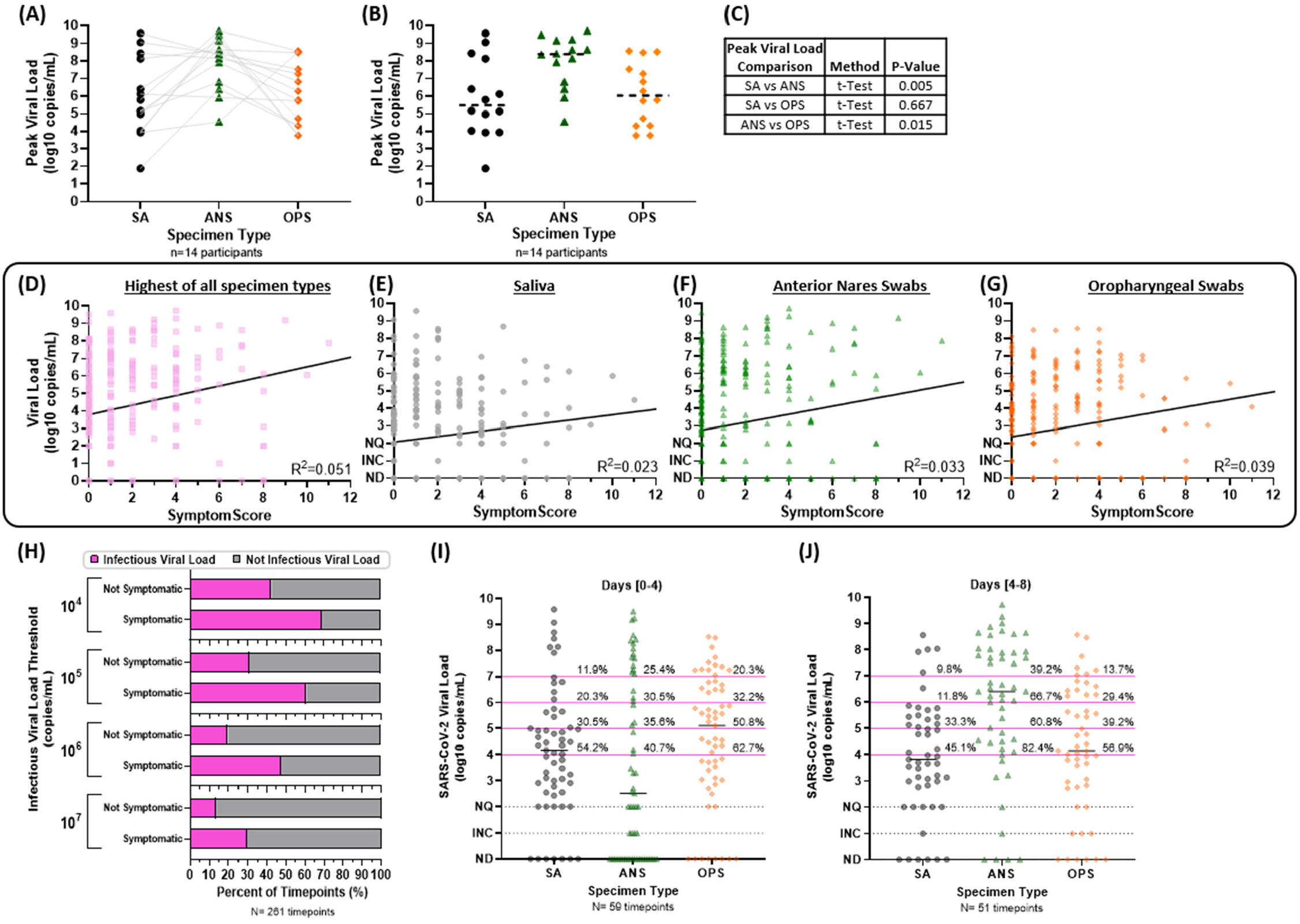
Peak and distribution of viral loads from the 14 participants enrolled before or at the incidence of acute SARS-CoV-2 infection. (**A)** The peak viral load for each participant is plotted with lines connecting to the viral loads of the other two specimen types at the same timepoint. **(B)** The distribution of peak viral loads for each specimen type is plotted; dashed horizontal bars indicate the medians. (**C**) Table showing statistical test results for comparisons of peak viral load in each specimen type, including the test method, performed in Graphpad Prism 9.2.0. For the cohort of 14 participants enrolled before or at the incidence of infection, the total number of symptoms reported at each timepoint was considered the Symptom Score. The Symptom Score was then plotted against the (**D**) highest viral load in all specimen types, the (**E**) viral load in SA specimens (**F**) ANS specimens and (**G**) OPS specimens. The text on each plot provides the Pearson correlation R squared value, and black lines indicate the line of best fit from linear regression. (**H**) For each symptomatic (Symptom Score >0) or asymptomatic timepoint, viral loads in any specimen type above the given IVLTs were considered infectious (magenta) and those below were considered not infectious (grey). The percentage of infectious and not infectious timepoints, for either symptomatic or not symptomatic timepoints is shown as a horizontal stacked bar graph. (**I**) The distribution of viral loads measured from a positive specimen of each specimen type during the first 4 days and (J) days 4 to 8 from the incidence of infection. N indicates the number of positive specimens of each type (by our high-analytical-sensitivity assay). Percentages above magenta lines to the right of each distribution indicate the fraction of all positive specimen of that type with a viral load at or above that infectious threshold. Black horizontal lines indicate the median viral load for each specimen type. SA, saliva; ANS, anterior-nares swab; OPS, oropharyngeal swab; NQ, below quantifiable; INC, inconclusive; ND, not detected.

**Figure S2.**
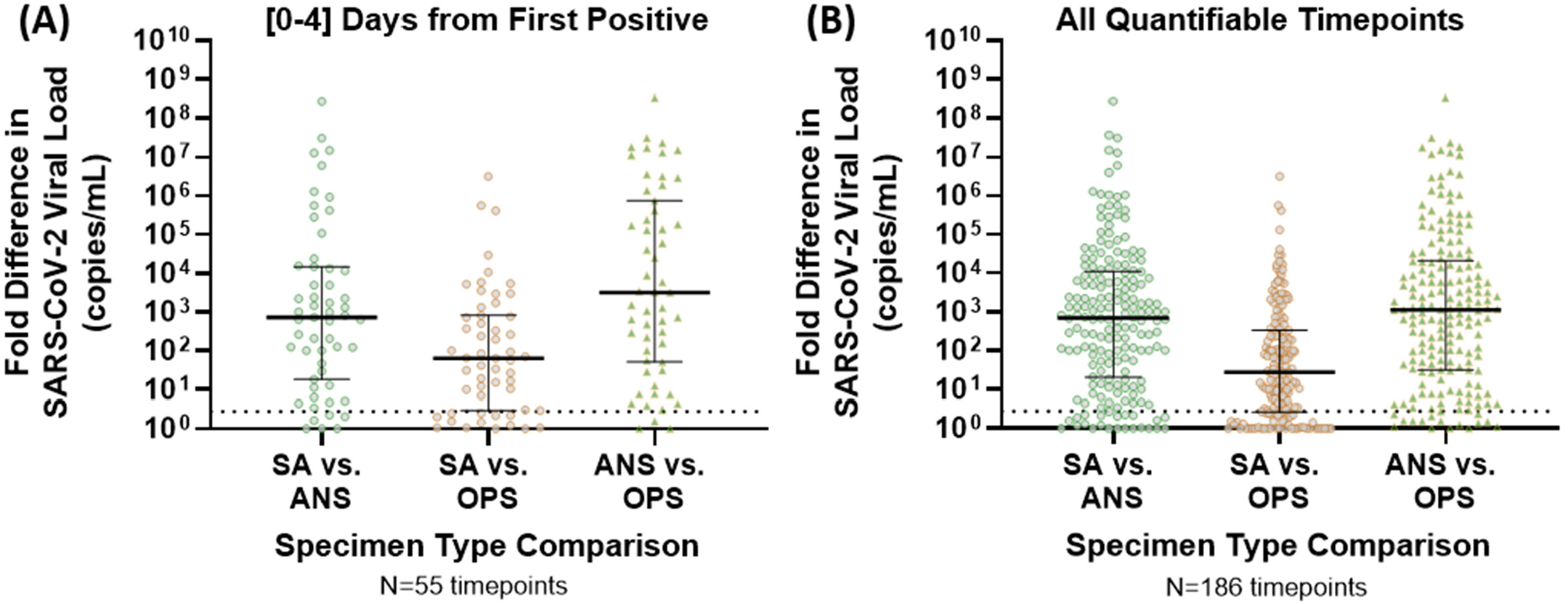
Relative (fold) differences in viral loads from paired specimen types. The fold difference (ratio of higher viral-load specimen of one type over a lower viral-load specimen of another type from the same participant at the same specimen-collection timepoint) are shown for **(A)** the first 4 days of infection (relative to first positive specimen of any type) and **(B)** for specimens collected at all timepoints when at least one specimen from the participant was positive for SARS-CoV-2. Specimens negative for SARS-CoV-2 or with viral loads below quantification had a viral load of 1 copy/mL imputed for calculations. Black bar indicates median. Dashed line indicates 2.8 fold difference, the level of *RNase P* sampling noise (**Fig S4**). SA, saliva; ANS, nasal anterior-nares swab; OPS. oropharyngeal swab, NQ indicates that both specimens being compared had unquantifiable viral loads so an absolute difference could not be calculated.

**Figure S3.**
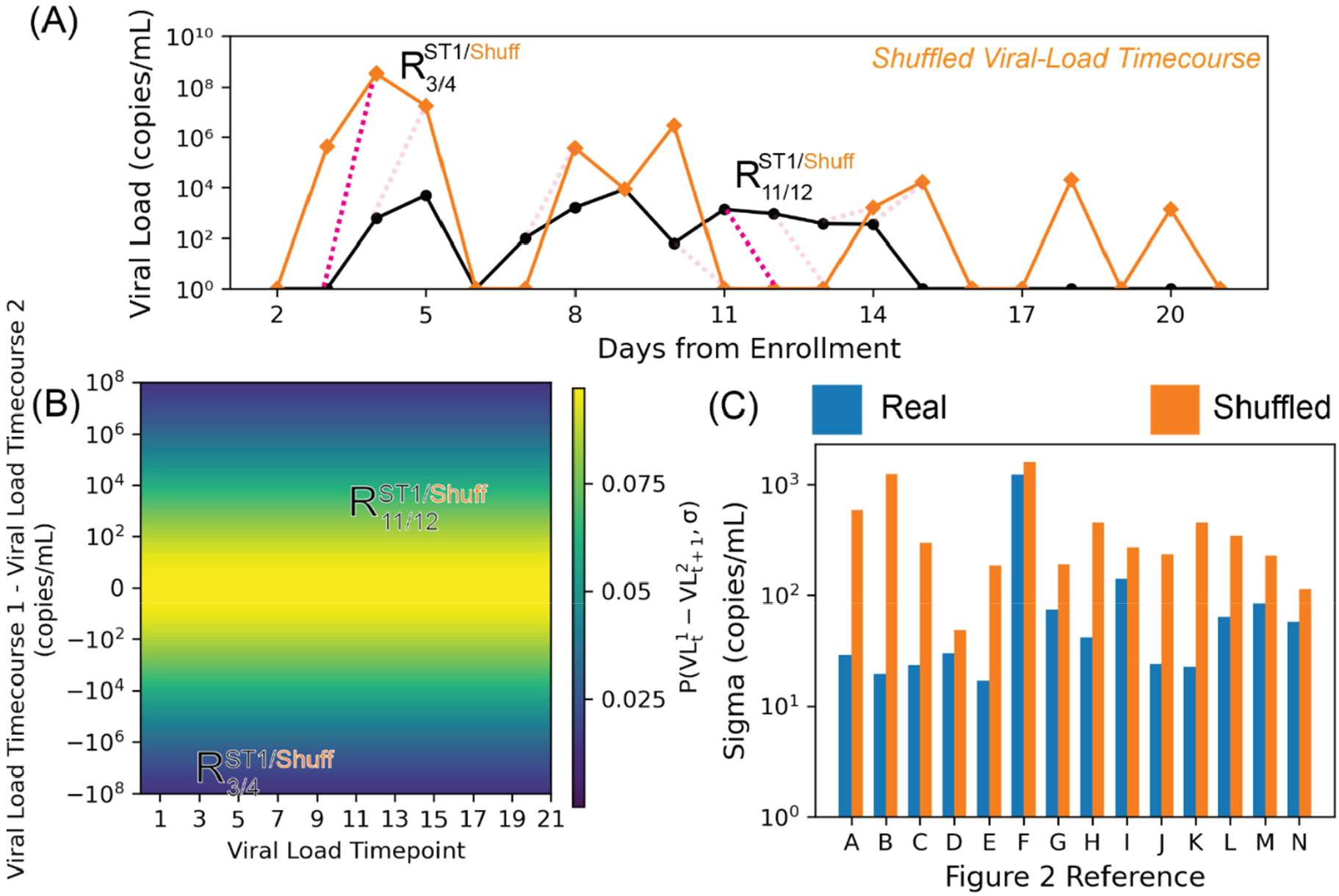
Increased Standard Deviation for Shuffled Viral-Load Timecourses Suggests Correct Sample Assignment by Participant and Specimen Type. **(A)** Viral-load timecourses for SA specimens collected from participant B (black). A “shuffled” timecourse (orange), obtained by randomizing specimen types at every timepoint, is shown in orange. This “shuffled” timecourse represents data that would be collected if an individual collected the incorrect specimen type when submitting samples. As in Figure S2, differences between timepoints for both “shuffled” and saliva timecourses were calculated. However, the timepoint after the one used for SA is selected for the “shuffled” timecourse. **(B)** Comparisons between pairwise differences between timepoints were visualized on a heatmap.. Background coloring represents the probability of observing pairwise residuals between the shuffled timecourse and the data from the saliva timecourse. Probabilities were generated from a normal distribution centered on 0 with a standard deviation (sigma) generated from the two timecourses. **(C)** Noise obtained from comparison of timecourses against themselves (blue) and shuffled equivalents (orange). Noise was estimated for each of the three specimen types for each individual. Estimates of noise from self-comparisons are statistically significantly from those obtained from comparisons with “shuffled timecourses” (*P*<0.001).

**Figure S4.**
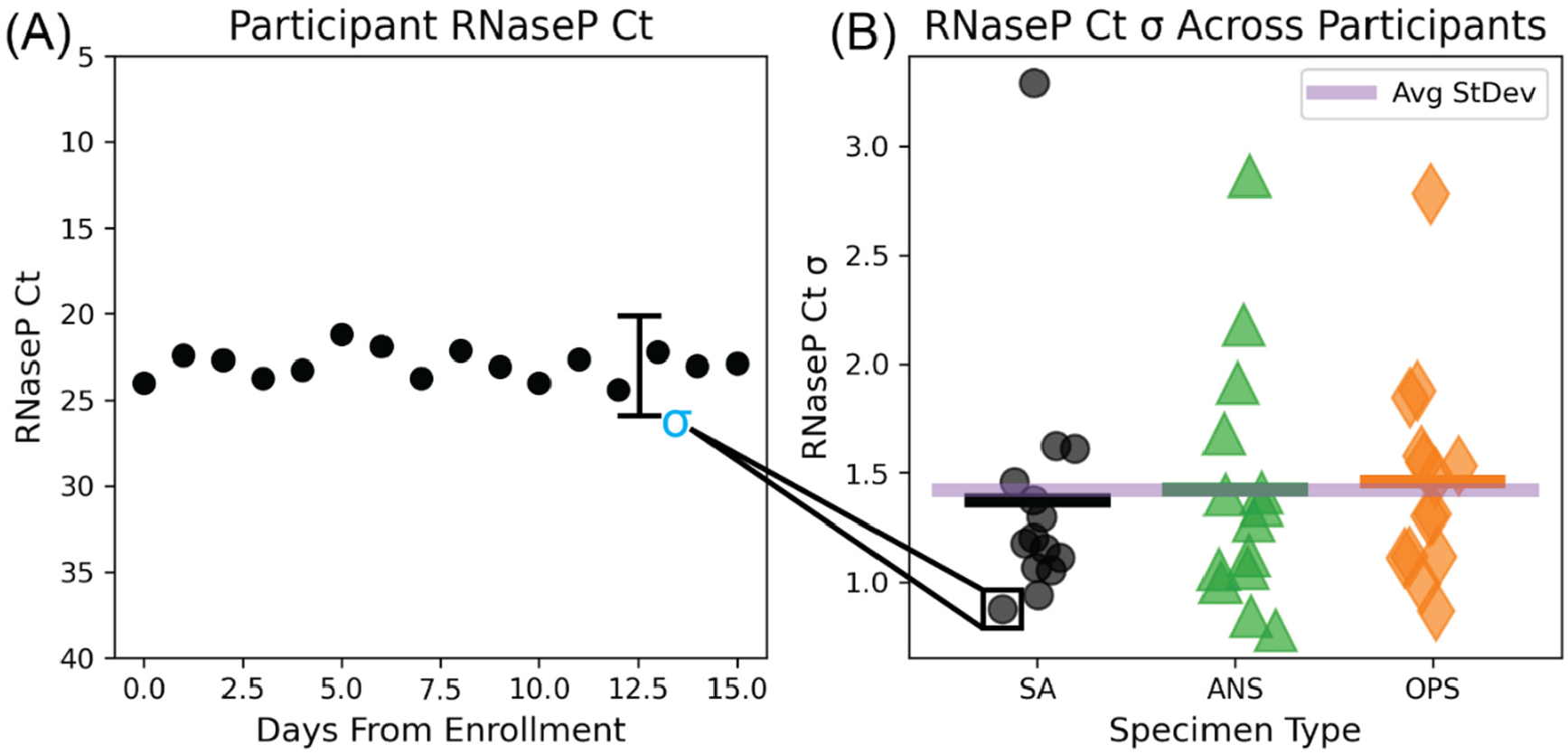
*RNase P* as a Measure of Sampling Variation for 14 Individuals Enrolled At or Before the Incidence of Infection. **(A)** Example longitudinal *RNase P* Ct measurements from a single individual. σ represents the standard deviation of the *RNase P* timecourse for a single individual in a single specimen type. (B) *RNase P* Ct standard deviations aggregated across specimen types and over all individuals. Horizontal black, green, and orange bars denote average standard deviations for each specimen type (saliva, SA; anterior-nares swab, ANS; oropharyngeal swab, OPS) across participants; the purple horizontal bar represents the average standard deviation over all participants and all specimen types.

**Figure S5.**
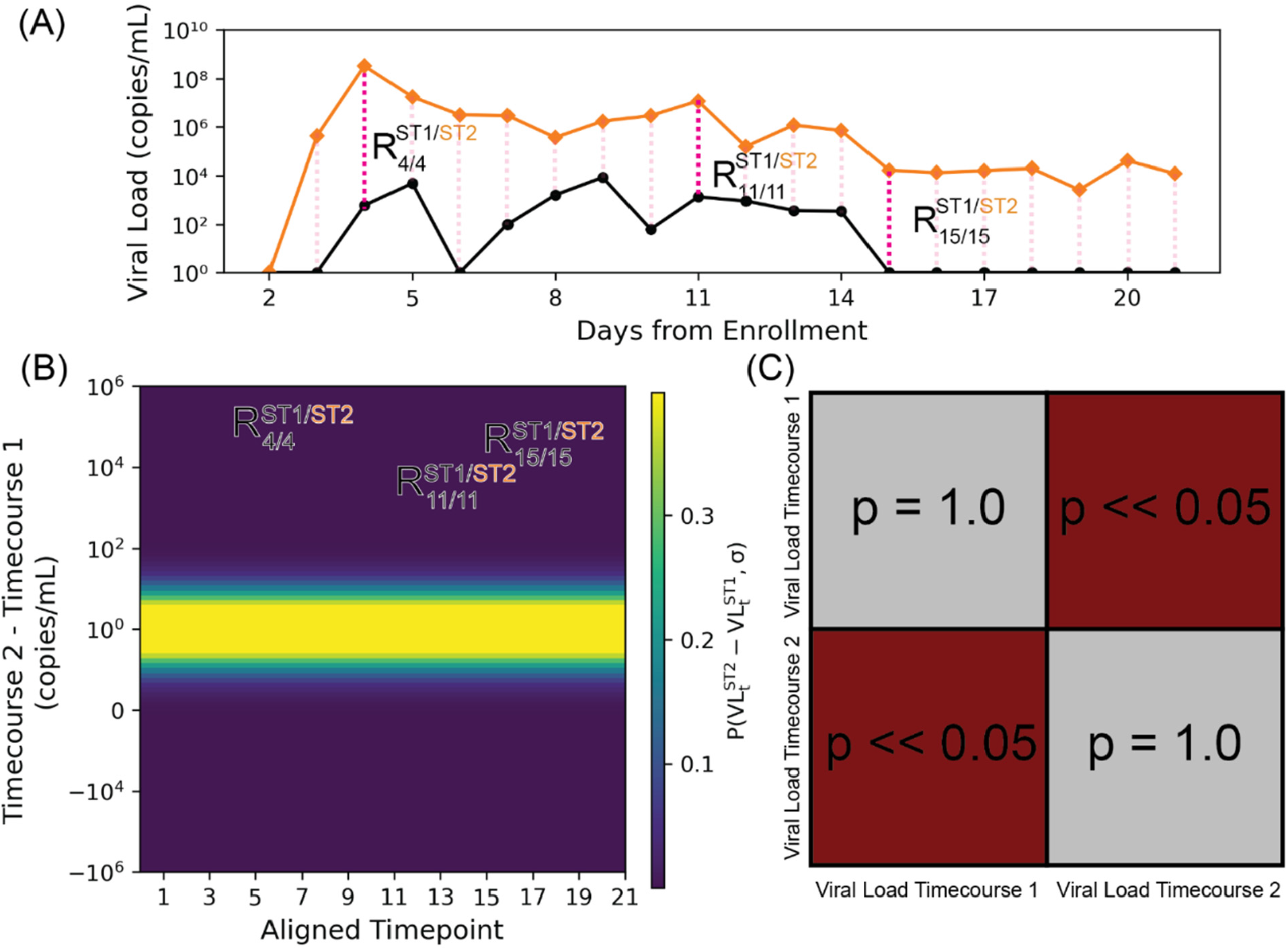
Pairwise Comparison of Viral-Load Timecourses. **(A)** As an example, the viral-load timecourses for saliva and oropharyngeal swab specimens collected from Z144 are shown. To compare two timecourses, first, the magnitude of the differences between the two timecourses at the same timepoint were calculated. Subscripts refer to time indices and superscripts refer to specimen types. **(B)** These differences were visualized on a graph with the x-axis representing the viral loads of the first timecourse and the y-axis representing the viral loads from the second timecourse. The line y=x, representing perfect agreement between the two timecourses, is plotted in red and background coloring represents probability of observing data given the null hypothesis that the two timecourses are equal. Such probabilities are either estimated from the timecourses themselves (Figure 3A) or from noise contained in *RNase P* data (Figure 3B). **(C)** Statistical significance of differences between viral-load timecourses. Absolute differences between timecourses were compared with the magnitude of bootstrapped noise samples and statistical significance was determined via an upper-tailed hypothesis test. Statistically significant timecourses are depicted in maroon and timecourses that are not significantly different are depicted in gray.

**Figure S6.**
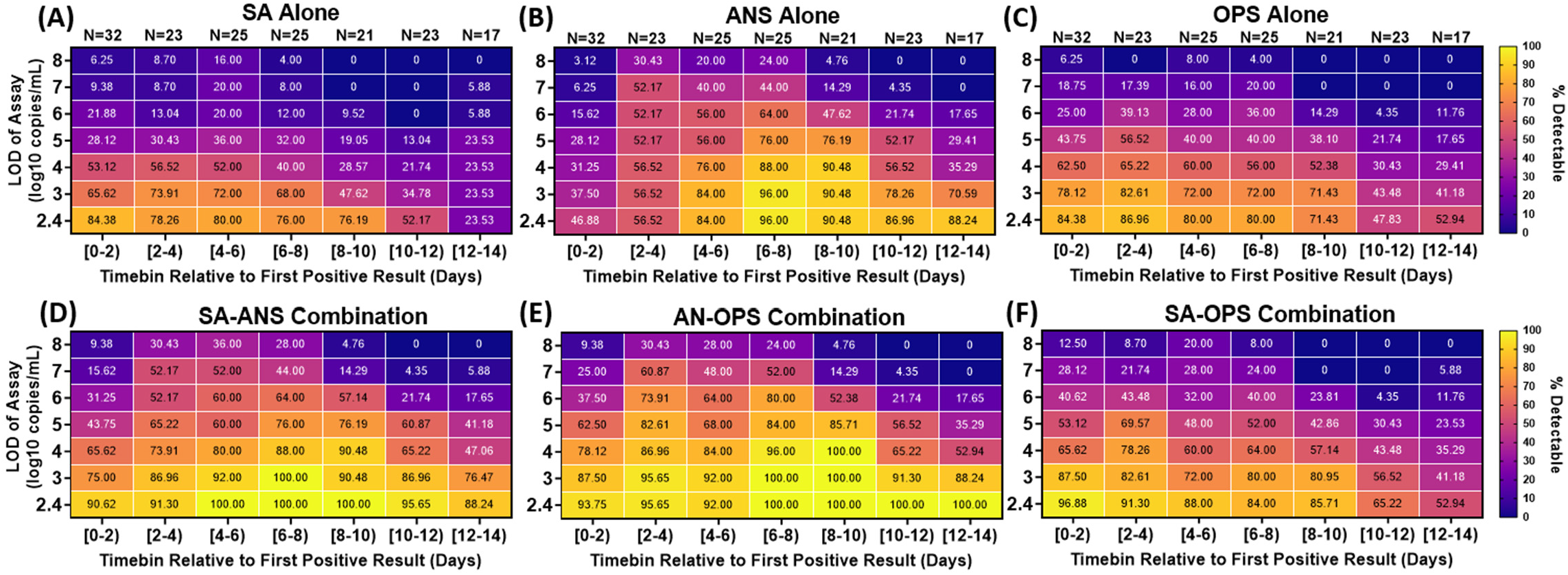
Extreme differences in viral load between specimen types result in low clinical sensitivity to detect infected persons by any single specimen type (A-C) but improved by combination specimen types (D-G). Heatmaps show the inferred clinical sensitivity for (**A**) saliva (SA) specimens alone (**B**) anterior-nares swab (ANS) specimens alone and (**C**) oropharyngeal swab (OPS) specimens alone, throughout the course of the infection (in two-day timebins relative to the first positive specimen of any type) for varying test LODs. Inferred clinical sensitivity was calculated as the number of specimens of the given type with viral loads greater than the given LOD, divided by the total number of specimens collected within that timebin. N indicates the number of specimens for each timebin. Only timepoints where at least one specimen had a quantifiable viral load (250 copies/mL) are included. (**D**) Inferred clinical sensitivity of a computationally-contrived specimen that combines saliva and anterior-nares swab (SA– ANS), **(E)** anterior-nares–oropharyngeal swab (AN–OPS) combination, **(F)** saliva and oropharyngeal swab (SA–OPS) combination, and (**G**) all three specimen types measured. The viral load for these contrived combination specimen types is the higher viral load from the specimen types included in the combination collected by a participant at a given timepoint. SA, saliva; ANS, anterior-nares swab; OPS, oropharyngeal swab. Four-day timebins are shown in **Fig 4**.

**Figure S7.**
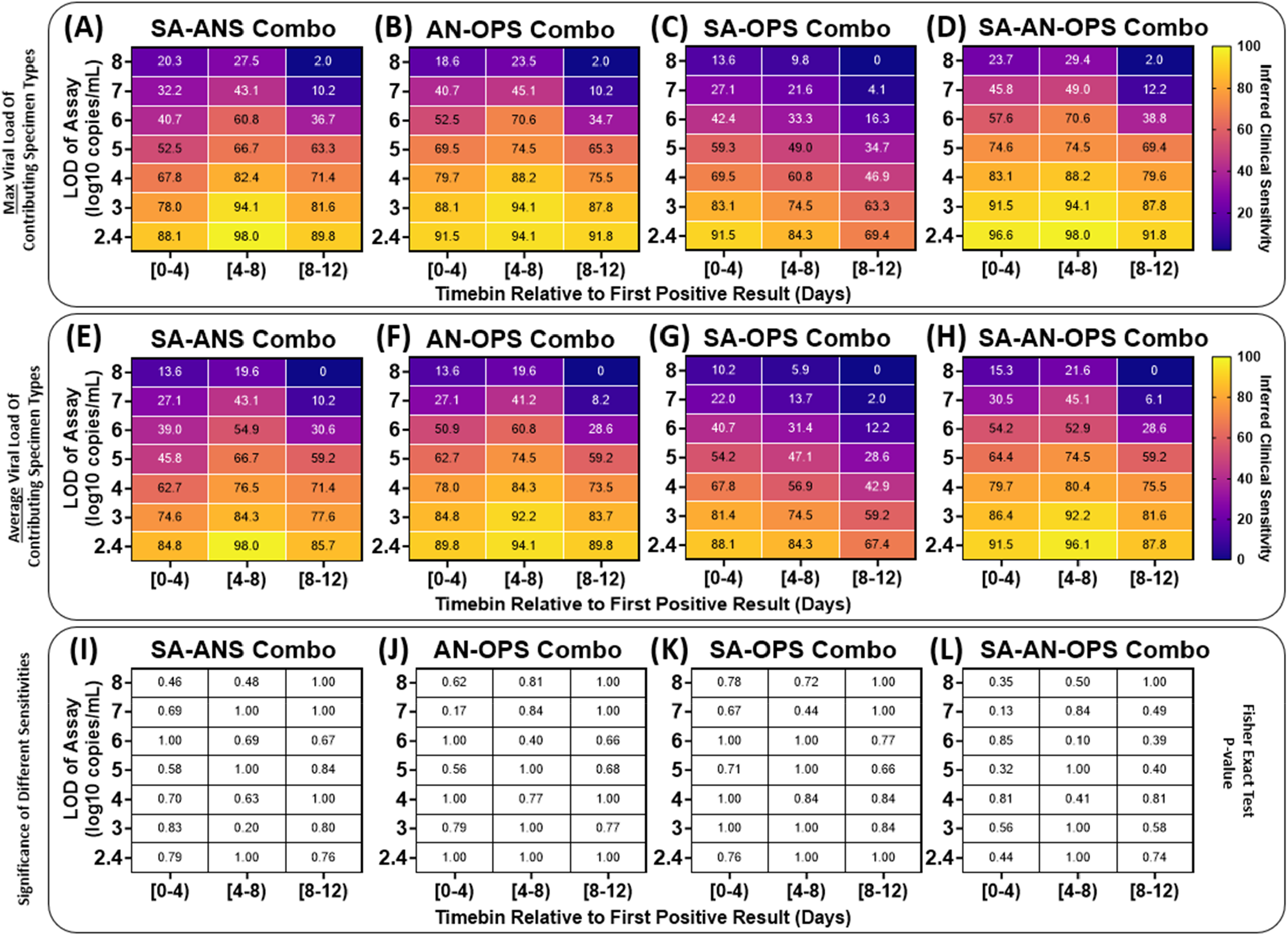
Inferred performance of computationally-contrived combination specimen types by averaging paired single specimen viral loads is similar to taking the maximum viral load of paired single specimen viral loads. Computationally-contrived combination specimen types were generated by taking a function of the viral loads from paired single specimen types collected by a participant at a timepoint. Detection of an infected person was inferred if the viral load in the computationally-contrived specimen type was above the LOD of the assay being used for testing (y-axis). The inferred clinical sensitivity of a given combination specimen type was calculated as the proportion of specimens inferred to be detectable at a given LOD over all positive specimen during each phase of the infection relative to the incidence of infection (x-axis), Each panel provides a heatmap colored by inferred clinical sensitivity when the viral load of computationally-contrived combination specimen types is calculated as the (**A-D**) maximum or (**E-H**) average viral load of paired single specimen types included in the combination, collected by a participant at a given timepoint. The binomial proportions using each function were compared with each other for each cell in each heatmap using the one-sided Fisher Exact Test with the alternative hypothesis that the maximum function would result in greater clinical sensitivity; resulting *P*-values are provided for respective cells in (**I-L**). SA– ANS, saliva–anterior-nares swab combination specimen: AN–OPS, anterior-nares–oropharyngeal swab combination specimen; SA–OPS, saliva– oropharyngeal swab combination specimen; SA–AN–OPS, saliva–anterior-nares–oropharyngeal swab combination specimen.

**Figure S8.**
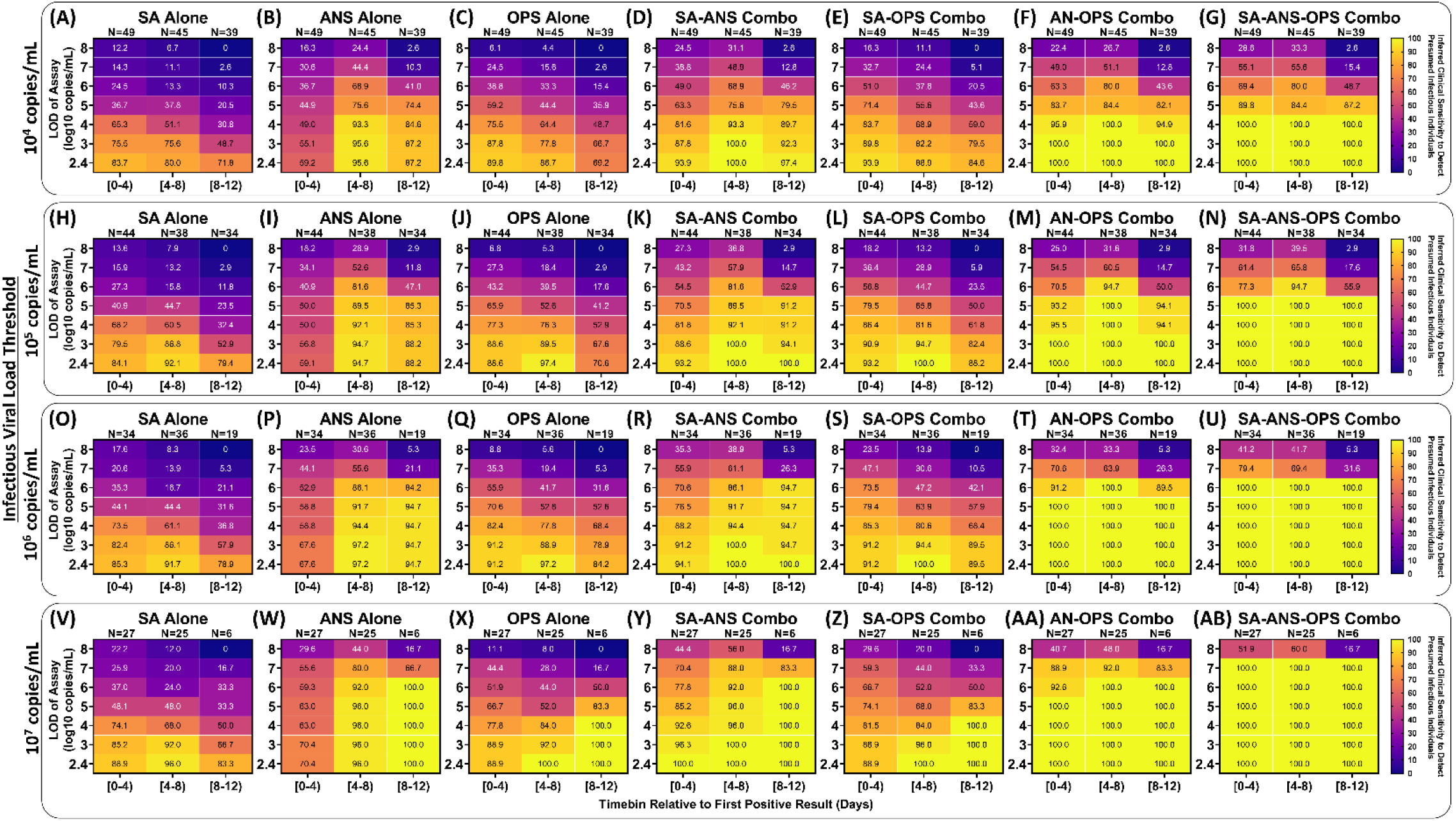
Inferred clinical sensitivity to detect presumed infectious individuals by testing single and combination specimen types using a range of test analytical sensitivities throughout acute, incident infection. For each 4-day timebin relative to the first SARS-CoV-2 positive specimen (of any type), participants were classified as being presumed infectious if viral load in any specimen type collected at a given timepoint was above an infectious viral load threshold (shown on the left side for each group of panels). The inferred clinical sensitivity of each specimen type to detect presumed infectious participants was calculated for each LOD as the number of specimens of that specimen type with a measured viral load at or above the LOD divided by the total specimen-collection timepoints included that timebin. The value inside each cell is the inferred clinical sensitivity to detect a presumed infectious person with that specimen type using an assay with the given LOD during that period of infection. The viral load of computationally-contrived combination specimen types was taken as the higher viral load of the specimen types included in the combination collected by a participant at a given timepoint. SA, saliva; ANS, anterior-nares swab; OPS, oropharyngeal swab. Two-day timebins are shown in **Fig S9**.

**Figure S9.**
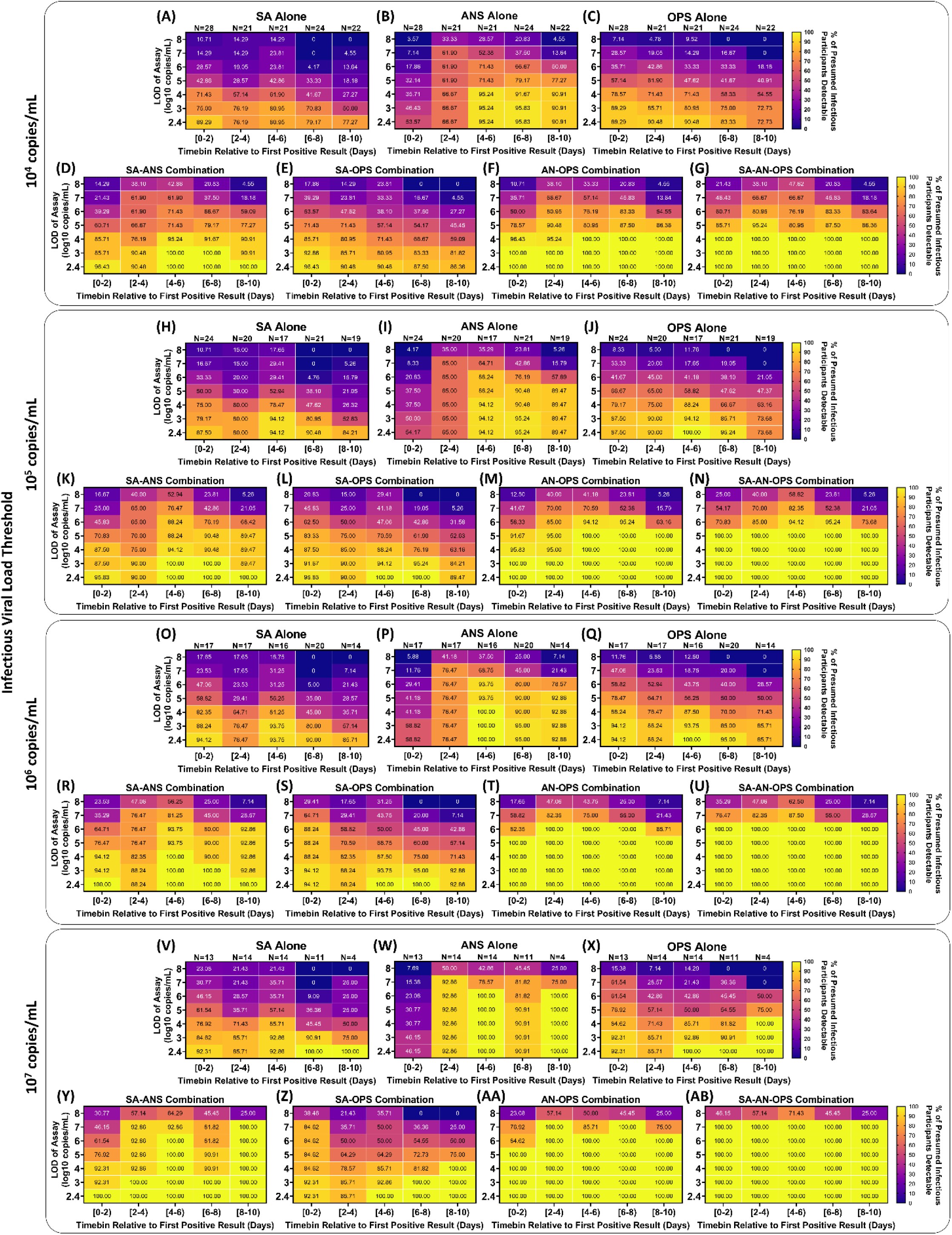
Inferred detection of presumed infectious individuals by single and combination specimen types and varying test analytical sensitivity throughout acute infection. For each two-day timebin relative to the first SARS-CoV-2 positive specimen (of any type), participants were classified as being presumed infectious based on whether the viral load in any specimen type collected at a given timepoint was above an infectious viral-load threshold (shown on the left side for each group of panels). The inferred clinical sensitivity of each specimen type to detect presumed infectious participants was calculated for each LOD as the number of specimens of that specimen type with a measured viral load at or above the LOD. The viral load of computationally-contrived combination specimen types was taken as the higher viral load of the specimen types included in the combination collected by a participant at a given timepoint. SA, saliva; ANS, anterior-nares swab; OPS, oropharyngeal swab. Four-day timebins are shown in **Fig S8**.

**Table S1.** Summary of the demographics, medical information, and vaccine history for the 14-participant cohort. Detailed information by participant can be found in Table S5.

| **<u>Sex\*</u>** |  |  |
| --- | --- | --- |
| Male | 7 | 50.0% |
| Female | 7 | 50.0% |
| **<u>Age</u>** |  |  |
| 6-11 | 2 | 14.3% |
| 12-17 | 1 | 7.1% |
| 18-29 | 2 | 14.3% |
| 30-39 | 3 | 21.4% |
| 40-49 | 4 | 28.6% |
| 50-59 | 2 | 14.3% |
| **<u>Race</u>** |  |  |
| White | 11 | 78.6% |
| Asian or Pacific Islander | 1 | 7.1% |
| Multiple Races | 2 | 14.3% |
| **<u>Ethnicity</u>** |  |  |
| Hispanic | 2 | 14.3% |
| Non-Hispanic | 12 | 85.7% |
| **<u>Tobacco Smoker or Vape User History</u>** |  |  |
| Current | 0 | 0.0% |
| Former | 2 | 14.3% |
| Never | 12 | 85.7% |
| **<u>Active Medications and Supplements</u>** |  |  |
| Vitamins/Supplements | 6 | 42.9% |
| Acetaminophen/NSAIDs | 3 | 21.4% |
| Allergy medications/Antihistamines | 2 | 14.3% |
| Antibiotics/Antivirals | 1 | 7.1% |
| **<u>Medical Comorbidities</u>** |  |  |
| Asthma | 1 | 7.1% |
| Anxiety or Depression | 2 | 14.3% |
| Diabetes | 1 | 7.1% |
| Overweight/Obesity | 6 | 42.9% |
| GI condition | 2 | 14.3% |
| **<u>SARS-CoV-2 Vaccination Status</u>** |  |  |
| Partially Vaccinated | 1 | 7.1% |
| Completed Vaccination | 5 | 35.7% |
| Fully vaccinated and boosted | 8 | 57.1% |
| No SARS-CoV-2 vaccines reported | 0 | 0.0% |

\*Participants were asked to report both sex at birth and current gender identity; all participants in this cohort responded cis-gender identities to sex at birth

**Table S2.** The Number of Presumed Infectious Specimens as a Factor of Specimen Type and Infectious Viral-Load Threshold.

| Specimen Type(s) | No. Presumed Infectious Specimens (%) by Infectious Viral Load |  |  |  |  |  |  |  |
| --- | --- | --- | --- | --- | --- | --- | --- | --- |
|  | 10<sup>4</sup> copies/mL |  | 10<sup>5</sup> copies/mL |  | 10<sup>6</sup> copies/mL |  | 10<sup>7</sup> copies/mL |  |
| SA Only | 7 | (4.4%) | 7 | (5.3%) | 6 | (6.5%) | 6 | (11%) |
| SA & NS | 6 | (3.9%) | 8 | (6%) | 5 | (5.4%) | 3 | (4.3%) |
| SA & OPS | 15 | (8.8%) | 8 | (6%) | 3 | (3.2%) | 1 | (2.9%) |
| SA & NS & OPS | 45 | (32%) | 24 | (20.7%) | 9 | (9.7%) | 3 | (4.3%) |
| NS & OPS | 12 | (11%) | 14 | (15%) | 12 | (12.9%) | 4 | (7.1%) |
| NS Only | 42 | (27%) | 43 | (31.3%) | 41 | (44.1%) | 29 | (51%) |
| OPS Only | 23 | (13%) | 21 | (16%) | 17 | (18.3%) | 13 | (19%) |
| **Total** |  | (100%) | 150 | (100%) | **93** | **(100%)** | 70 | (100%) |

**Table S3.**
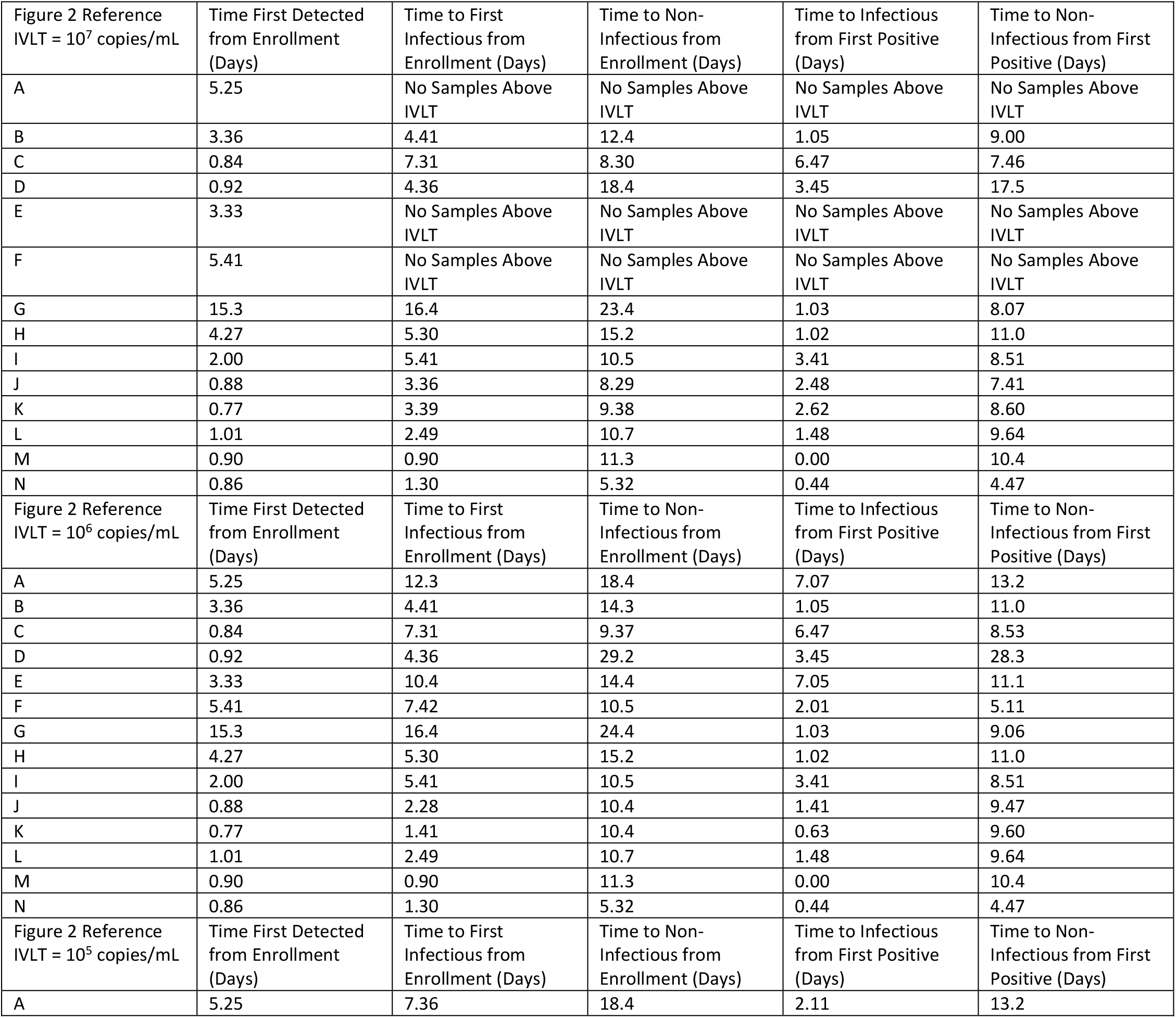
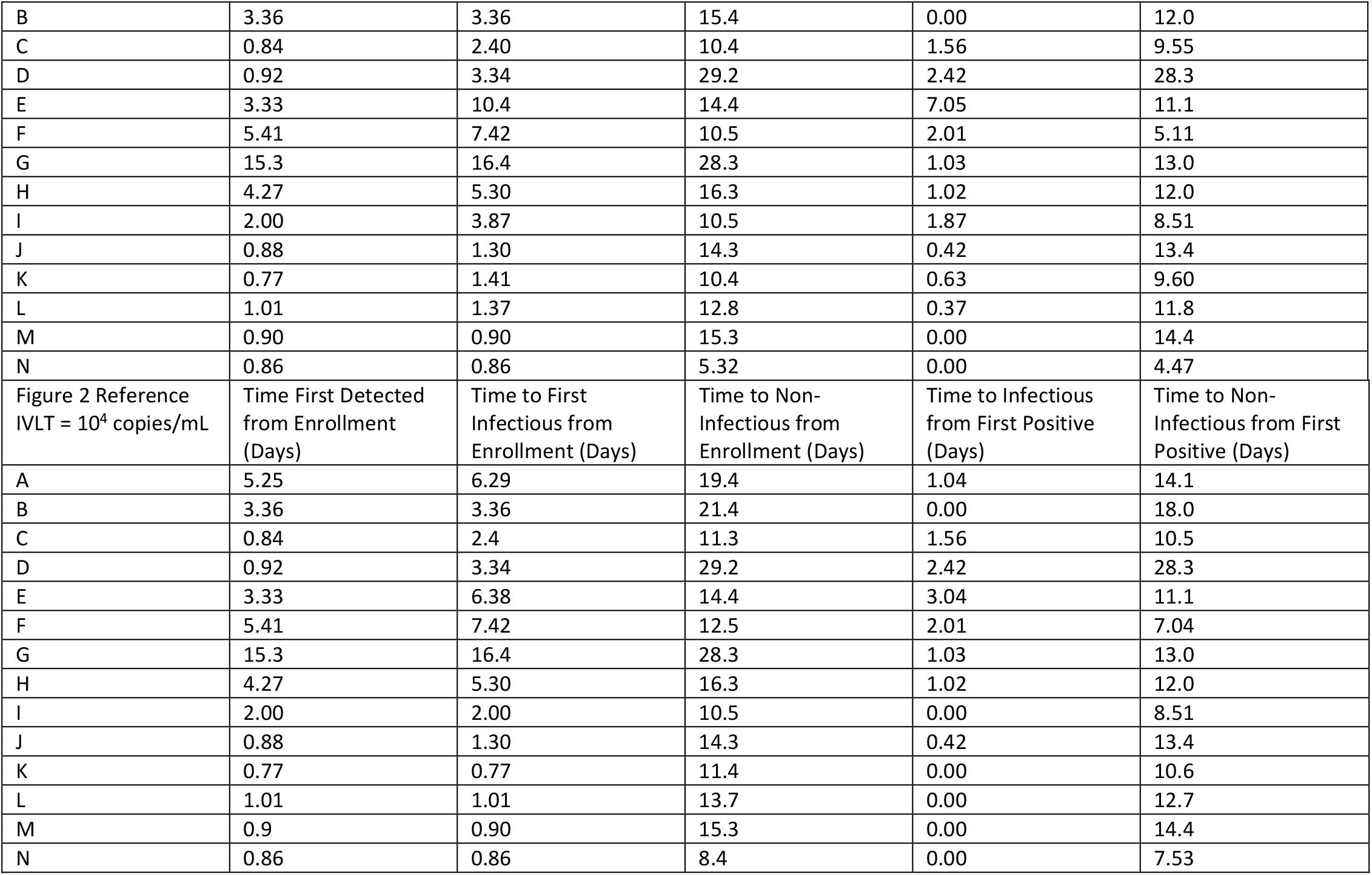
Times from First Positive by Any Specimen Type to First Viral Load Above Infectious Viral-Load Thresholds (IVLT) of 10^4^, 10^5^, 10^6^, 10^7^ copies/mL, and to First Timepoint with All Specimen Types Below IVLT.

| Figure 2 Reference<br>IVLT = 10<sup>7</sup> copies/mL | Time First Detected<br>from Enrollment<br>(Days) | Time to First<br>Infectious from<br>Enrollment (Days) | Time to Non-Infectious from<br>Enrollment (Days) | Time to Infectious<br>from First Positive<br>(Days) | Time to Non-Infectious from First<br>Positive (Days) |
| --- | --- | --- | --- | --- | --- |
| A | 5.25 | No Samples Above<br>IVLT | No Samples Above<br>IVLT | No Samples Above<br>IVLT | No Samples Above<br>IVLT |
| B | 3.36 | 4.41 | 12.4 | 1.05 | 9.00 |
| C | 0.84 | 7.31 | 8.30 | 6.47 | 7.46 |
| D | 0.92 | 4.36 | 18.4 | 3.45 | 17.5 |
| E | 3.33 | No Samples Above<br>IVLT | No Samples Above<br>IVLT | No Samples Above<br>IVLT | No Samples Above<br>IVLT |
| F | 5.41 | No Samples Above<br>IVLT | No Samples Above<br>IVLT | No Samples Above<br>IVLT | No Samples Above<br>IVLT |
| G | 15.3 | 16.4 | 23.4 | 1.03 | 8.07 |
| H | 4.27 | 5.30 | 15.2 | 1.02 | 11.0 |
| I | 2.00 | 5.41 | 10.5 | 3.41 | 8.51 |
| J | 0.88 | 3.36 | 8.29 | 2.48 | 7.41 |
| K | 0.77 | 3.39 | 9.38 | 2.62 | 8.60 |
| L | 1.01 | 2.49 | 10.7 | 1.48 | 9.64 |
| M | 0.90 | 0.90 | 11.3 | 0.00 | 10.4 |
| N | 0.86 | 1.30 | 5.32 | 0.44 | 4.47 |
| Figure 2 Reference<br>IVLT = 10<sup>6</sup> copies/mL | Time First Detected<br>from Enrollment<br>(Days) | Time to First<br>Infectious from<br>Enrollment (Days) | Time to Non-Infectious from<br>Enrollment (Days) | Time to Infectious<br>from First Positive<br>(Days) | Time to Non-Infectious from First<br>Positive (Days) |
| A | 5.25 | 12.3 | 18.4 | 7.07 | 13.2 |
| B | 3.36 | 4.41 | 14.3 | 1.05 | 11.0 |
| C | 0.84 | 7.31 | 9.37 | 6.47 | 8.53 |
| D | 0.92 | 4.36 | 29.2 | 3.45 | 28.3 |
| E | 3.33 | 10.4 | 14.4 | 7.05 | 11.1 |
| F | 5.41 | 7.42 | 10.5 | 2.01 | 5.11 |
| G | 15.3 | 16.4 | 24.4 | 1.03 | 9.06 |
| H | 4.27 | 5.30 | 15.2 | 1.02 | 11.0 |
| I | 2.00 | 5.41 | 10.5 | 3.41 | 8.51 |
| J | 0.88 | 2.28 | 10.4 | 1.41 | 9.47 |
| K | 0.77 | 1.41 | 10.4 | 0.63 | 9.60 |
| L | 1.01 | 2.49 | 10.7 | 1.48 | 9.64 |
| M | 0.90 | 0.90 | 11.3 | 0.00 | 10.4 |
| N | 0.86 | 1.30 | 5.32 | 0.44 | 4.47 |
| Figure 2 Reference<br>IVLT = 10<sup>5</sup> copies/mL | Time First Detected<br>from Enrollment<br>(Days) | Time to First<br>Infectious from<br>Enrollment (Days) | Time to Non-Infectious from<br>Enrollment (Days) | Time to Infectious<br>from First Positive<br>(Days) | Time to Non-Infectious from First<br>Positive (Days) |
| A | 5.25 | 7.36 | 18.4 | 2.11 | 13.2 |
| B | 3.36 | 3.36 | 15.4 | 0.00 | 12.0 |
| C | 0.84 | 2.40 | 10.4 | 1.56 | 9.55 |
| D | 0.92 | 3.34 | 29.2 | 2.42 | 28.3 |
| E | 3.33 | 10.4 | 14.4 | 7.05 | 11.1 |
| F | 5.41 | 7.42 | 10.5 | 2.01 | 5.11 |
| G | 15.3 | 16.4 | 28.3 | 1.03 | 13.0 |
| H | 4.27 | 5.30 | 16.3 | 1.02 | 12.0 |
| I | 2.00 | 3.87 | 10.5 | 1.87 | 8.51 |
| J | 0.88 | 1.30 | 14.3 | 0.42 | 13.4 |
| K | 0.77 | 1.41 | 10.4 | 0.63 | 9.60 |
| L | 1.01 | 1.37 | 12.8 | 0.37 | 11.8 |
| M | 0.90 | 0.90 | 15.3 | 0.00 | 14.4 |
| N | 0.86 | 0.86 | 5.32 | 0.00 | 4.47 |
| Figure 2 Reference<br>IVLT = 10<sup>4</sup> copies/mL | Time First Detected<br>from Enrollment<br>(Days) | Time to First<br>Infectious from<br>Enrollment (Days) | Time to Non-Infectious from<br>Enrollment (Days) | Time to Infectious<br>from First Positive<br>(Days) | Time to Non-Infectious from First<br>Positive (Days) |
| A | 5.25 | 6.29 | 19.4 | 1.04 | 14.1 |
| B | 3.36 | 3.36 | 21.4 | 0.00 | 18.0 |
| C | 0.84 | 2.4 | 11.3 | 1.56 | 10.5 |
| D | 0.92 | 3.34 | 29.2 | 2.42 | 28.3 |
| E | 3.33 | 6.38 | 14.4 | 3.04 | 11.1 |
| F | 5.41 | 7.42 | 12.5 | 2.01 | 7.04 |
| G | 15.3 | 16.4 | 28.3 | 1.03 | 13.0 |
| H | 4.27 | 5.30 | 16.3 | 1.02 | 12.0 |
| I | 2.00 | 2.00 | 10.5 | 0.00 | 8.51 |
| J | 0.88 | 1.30 | 14.3 | 0.42 | 13.4 |
| K | 0.77 | 0.77 | 11.4 | 0.00 | 10.6 |
| L | 1.01 | 1.01 | 13.7 | 0.00 | 12.7 |
| M | 0.9 | 0.90 | 15.3 | 0.00 | 14.4 |
| N | 0.86 | 0.86 | 8.4 | 0.00 | 7.53 |

**Table S4.** Statistical comparisons of inferred clinical sensitivity drawn from Fig 7 and 8. For select comparisons (across specimen types, assay LODs, infection stages/timebins, or IVLTs), the comparison is stated, along with the inferred clinical sensitivity (with 95% Confidence Intervals), statistical method, and significance of the difference. Index is referenced in the main text. Bolded cells in each row indicate the groups being compared. Values under Contingency Table indicate number of specimens. ‘Infectious’ indicates timepoints from individuals with a viral load in any specimen type above the infectious viral-load threshold listed in parentheses. Test Methods: A- Lower-Tailed McNemar Exact Test, B- Upper-Tailed McNemar Exact Test, C- Two-Tailed McNemar Exact Test, D- Lower-Tailed Fisher Exact Test. SA, saliva; ANS, anterior-nares swab; OPS, oropharyngeal swab; AN–OP, anterior-nares–oropharyngeal combination swab; SA–ANS, saliva–anterior-nares combination specimen; SA–OPS, saliva–oropharyngeal swab combination specimen; SA–ANS–OPS, saliva–anterior-nares–oropharyngeal swab combination specimen.

**Table S5.** Demographic and Medical Information for the Participants Shown in Fig 3. SARS-CoV-2 variant was determined by ANS swab in all cases except individual (B) who had low ANS viral loads so viral load was sequenced from a throat swab. The variant for participant (I) is inferred from the household index case.

| Fig 3 panel | Status on enrollment |  |  | Months* since vaccine # |  |  | Active Medications | Comorbidities/ medical conditions | Gender** | Age range (in years) | Race | Ethnicity | SARS-CoV-2 Variant |
| --- | --- | --- | --- | --- | --- | --- | --- | --- | --- | --- | --- | --- | --- |
|  | Saliva PCR | Throat PCR | Nasal PCR | 1st dose | 2nd dose | 3rd dose |  |  |  |  |  |  |  |
| (A) | neg | neg | neg | 9 [M] | 8 [M] | <2 [M] | n/a | n/a | male | 40-49 | White | not Hispanic | Omicron BA.1.1 |
| (B) | neg | neg | neg | 11 [JJ] | 3 [P] | none | PPI, vitamin/ supplement | obesity, GI condition, anxiety or depression | female | 30-39 | White | not Hispanic | Omicron BA.1.1 |
| (C) | inc | neg | neg | <1 [P] | none | none | acetaminophen | n/a | male | 6-11 | Multiple Races | not Hispanic | Omicron BA.1.1 |
| (D) | neg | neg | neg | 10 [M] | 9 [M] | 2 [M] | none | obesity | male | 30-39 | Asian or Pacific Islander | not Hispanic | Omicron BA.1.1 |
| (E) | neg | neg | neg | >11 [P] | <10 [P] | <3 [P] | allergy medication; acetaminophen, antihistamine, dextromethorphan, phenylephrine HCl, doxylamine | obesity | female | 30-39 | White | Hispanic | Omicron BA.1 |
| (F) | neg | neg | neg | 10 [P] | 9 [P] | none | vitamin/ supplement | n/a | female | 18-29 | White | not Hispanic | Omicron BA.1.1 |
| (G) | neg | neg | neg | <2 [P] | <1 [P] | none | vitamin/ supplement | n/a | male | 6-11 | White | not Hispanic | Omicron BA.1.1 |
| (H) | neg | neg | neg | 10 [M] | 9 [M] | 2 [M] | vitamin/ supplement | n/a | female | 40-49 | White | not Hispanic | Omicron BA.1.1 |
| (I) | neg | neg | neg | 10 [P] | 9 [P] | none | antibiotic, vitamin/ supplement | obesity | male | 18-29 | White | Hispanic | Omicron BA.1.1 (index case) |
| (J) | pos | pos | inc | 9 [M] | 8 [M] | <2 [M] | vitamin/ supplement | anxiety or depression | female | 40-49 | White | not Hispanic | Omicron BA.1.1 |
| (K) | pos | pos | inc | 9.5 [M] | 8.5 [M] | 0.5 [P] | NSAID | n/a | male | 40-49 | White | not Hispanic | Omicron BA.1.1 |
| (L) | pos | pos | pos | 11 [P] | 10 [P] | 2 [P] | allergy medication, diabetes medication, cholesterol medication | diabetes, high blood pressure, obesity, asthma, sleep apnea, GI condition | female | 50-59 | Multiple Races | not Hispanic | Omicron BA.1.1 |
| (M) | pos | pos | neg | 10 [M] | 9 [M] | 2 [M] | SSRI | oveweight, anxiety or depression | male | 50-59 | White | not Hispanic | Omicron BA.1.1 |
| (N) | pos | neg | pos | 5 [P] | 4[P] | none | none | n/a | female | 12-17 | White | not Hispanic | Omicron BA.1.1 |

\* Months from vaccine date are given relative to enrollment date

## AUTHOR CONTRIBUTIONS (listed alphabetically by last name)

Reid Akana (RA): Collaborated with AVW in creating digital participant symptom surveys; assisted with data quality control/curation with NS, HD, SC; created current laboratory information management system (LIMS) for specimen logging and tracking. Creation of iOS application for sample logging/tracking. Configured an SQL database for data storage. Created an Apache server and websites to view study data. Configured FTPS server to catalog PCR data. Wrote a Python package to access study data. Trained study coordinators on SQL. Troubleshooting and QC of LIMS. Made Fig 3(C-D) and 5D, and SI Figs S3, S4, S5, Table S2, S3, S4, S5, S6. Wrote and edited the manuscript with AVW and NS.

Alyssa M. Carter (AMC): Assisted with the inventory and archiving of >6,000 samples at Caltech; coordinated shipment of samples to Caltech with AER and JRBR; assisted with procurement of antigen tests; assisted with organizing volunteers and making participant kits; assisted AER in developing and implementing QC for participant kits. Provided feedback and edited the manuscript.

Yap Ching Chew (YCC): Primary liaison with Caltech team. Prepared and provided Zymo SafeCollect kits and related materials to Caltech team. Supervised the extraction, PCR, and QC teams at Pangea Laboratory. Sent PCR results daily to Caltech team. Arranged for Pangea team to perform viral-variant sequencing on selected samples; reported results and provided sequencing files.

Saharai Caldera (SC): Study coordinator; recruited, enrolled and maintained study participants with NS and HD; study-data quality control, curation and archiving with RA, NS, HD and MKK; supplies acquisition with AER, NS, HD and MKK.

Hannah Davich (HD): Lead study coordinator; co-wrote participant informational sheets with NS; developed recruitment strategies and did outreach with NS; participant kit creation and co-coordinated kit-making by volunteers with AER; recruited, enrolled and maintained study participants with NS and SC; managed the study-coordinator inventory; study-data quality control, curation and archiving with RA, NS, SC and MKK; supplies acquisition with AER, NS, SC and MKK.

Matthew Feaster (MF): Co-investigator; collaborated with AVW, MMC, NS, YG, RFI on study design and recruitment strategies; provided guidance and expertise on SARS-CoV-2 epidemiology and local trends.

Ying-Ying Goh (Y-YG): Co-investigator; collaborated with AVW, MMC, NS, MF, RFI on study design and recruitment strategies; provided guidance and expertise on SARS-CoV-2 epidemiology and local trends.

Rustem F. Ismagilov (RFI): Principal investigator; collaborated with AVW, MMC, NS, MF, YYG on study design and recruitment strategies; provided leadership, technical guidance, oversight of all analyses, and was responsible for obtaining the primary funding for the study.

Mi Kyung Kim (MKK): Study coordinator (part-time); maintaining participants with NS, HD, and SC; study-data quality control, curation and archiving with RA, NS, SC and HD; supplies acquisition with AER, NS, SC and HD; collected contact info for local university/college student health centers for recruitment outreach; assembled Table S5 with NS.

John Raymond B. Reyna (JRBR): Organized sample labeling and short-term storage of all samples at Pangea Laboratories. Arranged shipment of all samples to Caltech team. Assisted with processing of the specimens.

Anna E. Romano (AER): Co-coordinated kit-making by volunteers with HD; implemented QC process for kit-making; participated in kit-making; managed logistics for the inventory and archiving of >6,000 samples at Caltech; supplies acquisition with HD, NS, SC and MKK; assisted with securing funding; compiled Table S3; organized and performed QC on sequencing data. Provided feedback and edited the manuscript.

Natasha Shelby (NS): Study administrator; collaborated with AVW, RFI, YG, MF on initial study design and recruitment strategies; co-wrote IRB protocol and informed consent with AVW; co-wrote enrollment questionnaire and post-study questionnaire with AVW; initiated the collaboration with Zymo and served as primary liaison throughout study; reviewed pilot sampling data and amended instructional sheets/graphics for specimen collections in collaboration with Zymo; co-wrote participant informational sheets with HD; hired, trained, and supervised the study-coordinator team; developed recruitment strategies and did outreach with HD; recruited, enrolled and maintained study participants with HD and SC; co-developed participant keep/drop criteria with AVW; performed the daily upload, review, and QC of PCR data received from Zymo; made the daily keep/drop decisions based on viral-load trajectories in each household; made all phone calls to alert presumptive positives of their status and provide resources; study-data quality control, curation and archiving with RA, HD, SC and MKK; organized archiving of all participant data and antigen-test photographs; supplies acquisition with AER, HD, SC and MKK; assisted with securing funding; managed the overall study budget; assembled Figs 1-2 with AVW; assembled Table S1; assembled Table S5 with MKK; managed citations and reference library; verified the underlying data with AVW and RA; co-wrote and edited the manuscript with AVW and RA.

Matt Thomson (MT): Assisted with statistical approach and analyses.

Colten Tognazzini (CT): Coordinated the recruitment efforts at PPHD with case investigators and contact tracers; provided guidance and expertise on SARS-CoV-2 epidemiology and local trends.

Alexander Viloria Winnett (AVW): Collaborated with NS, RFI, YG, MF on initial study design and recruitment strategies; co-wrote IRB protocol and informed consent with NS; co-wrote enrollment questionnaire and post-study questionnaire with NS; co-developed participant keep/drop criteria with NS; funding acquisition; designed and coordinated LOD validation experiments; selected and prepared specimen for viral-variant sequencing with NS, YC, and AER; assisted with the inventory and archiving of >6,000 specimen at Caltech with AER and AMC; minor role supporting outreach by HD and NS; minor role supporting kit-making by AER, HD and AMC; verified the underlying data with NS and RA; assembled Figs 1-2 with NS; performed analysis and prepared Figs 4-7, Table S2, Fig S1, S2, S6, S7, S8, S9. Co-wrote and edited the manuscript with NS and RA.

Taikun Yamada (TY): Performed the RT-qPCR COVID-19 testing at Pangea Laboratory.

## References

1. Elie B, et al. (2022) Variant-specific SARS-CoV-2 within-host kinetics. J. Med. Virol. 94(8):3625–3633.

2. Laajaj R, et al. (2022) Understanding how socioeconomic inequalities drive inequalities in COVID-19 infections. Sci. Rep. 12(1):8269.

3. Hart WS, Maini PK, & Thompson RN (2021) High infectiousness immediately before COVID-19 symptom onset highlights the importance of continued contact tracing. eLife 10:e65534.

4. NIH (2021) Therapeutic Management of Adults With COVID-19. https://www.covid19treatmentguidelines.nih.gov/therapeutic-management/

5. Beigel JH, et al. (2020) Remdesivir for the Treatment of Covid-19 — Final Report. N. Engl. J. Med. 383(19):1813–1826.

6. Libster R, et al. (2020) Prevention of severe COVID-19 in the elderly by early high-titer plasma. medRxiv:2020.2011.2020.20234013.

7. Kim Y-g, et al. (2017) Comparison between Saliva and Nasopharyngeal Swab Specimens for Detection of Respiratory Viruses by Multiplex Reverse Transcription-PCR. J. Clin. Microbiol. 55(1):226.

8. Liu J, et al. (2021) SARS-CoV-2 cell tropism and multiorgan infection. Cell Discovery 7(1):17.

9. Huang N, et al. (2021) SARS-CoV-2 infection of the oral cavity and saliva. Nat. Med. 27(5):892–903.

10. Hanson KE, et al. (2021) The Infectious Diseases Society of America Guidelines on the Diagnosis of COVID-19: Molecular Diagnostic Testing. Clin. Infect. Dis. doi: 10.1093/cid/ciab048

11. Marais G, et al. (2021) Saliva swabs are the preferred sample for Omicron detection. medRxiv:2021.2012.2022.21268246.

12. Chu CY, et al. (2021) Performance of saliva and mid-turbinate swabs for detection of the beta variant in South Africa. The Lancet Infectious Diseases 21(10):1354.

13. Savela ES, et al. (2021) Quantitative SARS-CoV-2 viral-load curves in paired saliva and nasal swabs inform appropriate respiratory sampling site and analytical test sensitivity required for earliest viral detection. J. Clin. Microbiol. 0(ja):JCM.01785-01721.

14. Lin J, et al. (2022) Where is Omicron? Comparison of SARS-CoV-2 RT-PCR and Antigen Test Sensitivity at Commonly Sampled Anatomic Sites Over the Course of Disease. medRxiv:2022.2002.2008.22270685.

15. Galliez Rafael M, et al. (Evaluation of the Panbio COVID-19 Antigen Rapid Diagnostic Test in Subjects Infected with Omicron Using Different Specimens. Microbiology Spectrum 0(0):e01250–01222.

16. Killingley B, et al. (2022) Safety, tolerability and viral kinetics during SARS-CoV-2 human challenge in young adults. Nat. Med. 28(5):1031–1041.

17. Adamson B, Sikka R, Wyllie AL, & Premsrirut P (2022) Discordant SARS-CoV-2 PCR and Rapid Antigen Test Results When Infectious: A December 2021 Occupational Case Series. medRxiv:2022.2001.2004.22268770.

18. Binnicker MJ (2022) Should I Use a Throat Swab for My At-Home COVID-19 Antigen Test? in Forbes. https://www.forbes.com/sites/coronavirusfrontlines/2022/01/25/should-i-use-a-throat-swab-for-my-at-home-covid-19-antigen-test/?sh=5adc7f3b3088

19. Arnaout R, et al. (2021) The Limit of Detection Matters: The Case for Benchmarking Severe Acute Respiratory Syndrome Coronavirus 2 Testing. Clin. Infect. Dis. 73(9):e3042–e3046.

20. Winnett A, et al. (2020) SARS-CoV-2 Viral Load in Saliva Rises Gradually and to Moderate Levels in Some Humans. medRxiv:2020.2012.2009.20239467.

21. Lai J, et al. (2022) Comparison of Saliva and Mid-Turbinate Swabs for Detection of COVID-19. medRxiv:2021.2012.2001.21267147.

22. European Centre for Disease Prevention and Control (2022) Diagnostic Testing and Screening for SARS-CoV-2. https://www.ecdc.europa.eu/en/covid-19/latest-evidence/diagnostic-testing

23. Drain PK (2022) Rapid Diagnostic Testing for SARS-CoV-2. N. Engl. J. Med. 386(3):264–272.

24. Rader B, et al. (2022) Use of At-Home COVID-19 Tests - United States, August 23, 2021-March 12, 2022. MMWR Morb. Mortal. Wkly. Rep. 71(13):489–494.

25. Wölfel R, et al. (2020) Virological assessment of hospitalized patients with COVID-2019. Nature 581(7809):465–469.

26. Ke R, et al. (2022) Daily longitudinal sampling of SARS-CoV-2 infection reveals substantial heterogeneity in infectiousness. Nature Microbiology 7(5):640–652.

27. Walsh KA, et al. (2020) SARS-CoV-2 detection, viral load and infectivity over the course of an infection. The Journal of infection 81(3):357–371.

28. Rhoads DD & Pinsky BA (2021) The Truth about SARS-CoV-2 Cycle Threshold Values Is Rarely Pure and Never Simple. Clin. Chem. 68(1):16–18.

29. Poon KS & Wen-Sim Tee N (2021) Caveats of Reporting Cycle Threshold Values from Severe Acute Respiratory Syndrome Coronavirus 2 (SARS-CoV-2) Qualitative Polymerase Chain Reaction Assays: A Molecular Diagnostic Laboratory Perspective. Clin. Infect. Dis. 73(9):e2851–e2852.

30. van Kampen JJA, et al. (2021) Duration and key determinants of infectious virus shedding in hospitalized patients with coronavirus disease-2019 (COVID-19). Nat Commun 12(1):267.

31. Perera R, et al. (2020) SARS-CoV-2 Virus Culture and Subgenomic RNA for Respiratory Specimens from Patients with Mild Coronavirus Disease. Emerg. Infect. Dis. 26(11):2701–2704.

32. Pickering S, et al. (2021) Comparative performance of SARS-CoV-2 lateral flow antigen tests and association with detection of infectious virus in clinical specimens: a single-centre laboratory evaluation study. Lancet Microbe 2(9):e461–e471.

33. Perera RAPM, et al. (2020) SARS-CoV-2 virus culture from the upper respiratory tract: Correlation with viral load, subgenomic viral RNA and duration of illness. medRxiv:2020.2007.2008.20148783.

34. L’Huillier AG, Torriani G, Pigny F, Kaiser L, & Eckerle I (2020) Shedding of infectious SARS-CoV-2 in symptomatic neonates, children and adolescents. medRxiv:2020.2004.2027.20076778.

35. Jones Terry C, et al. (2021) Estimating infectiousness throughout SARS-CoV-2 infection course. Science 373(6551):eabi5273.

36. Quicke K, et al. (2020) Longitudinal Surveillance for SARS-CoV-2 RNA Among Asymptomatic Staff in Five Colorado Skilled Nursing Facilities: Epidemiologic, Virologic and Sequence Analysis. medRxiv:2020.2006.2008.20125989.

37. Puhach O, et al. (2022) Infectious viral load in unvaccinated and vaccinated patients infected with SARS-CoV-2 WT, Delta and Omicron. medRxiv:2022.2001.2010.22269010.

38. Bal A, et al. (2020) Clinical and microbiological assessments of COVID-19 in healthcare workers: a prospective longitudinal study. medRxiv:2020.2011.2004.20225862.

39. Holmdahl I, Kahn R, Hay J, Buckee CO, & Mina M (2021) Estimation of Transmission of COVID-19 in Simulated Nursing Homes With Frequent Testing and Immunity-Based Staffing. JAMA Network Open 4(5):e2110071.

40. Schuit E, et al. (2021) Diagnostic accuracy of rapid antigen tests in asymptomatic and presymptomatic close contacts of individuals with confirmed SARS-CoV-2 infection: cross sectional study. BMJ 374:n1676.

41. Lin Y, et al. (2022) Incorporating temporal distribution of population-level viral load enables real-time estimation of COVID-19 transmission. Nat Commun 13(1):1155.

42. Goyal A, Reeves DB, Cardozo-Ojeda EF, Schiffer JT, & Mayer BT (2021) Viral load and contact heterogeneity predict SARS-CoV-2 transmission and super-spreading events. Elife 10.

43. Gomez J, Prieto J, Leon E, & Rodríguez A (2021) INFEKTA—An agent-based model for transmission of infectious diseases: The COVID-19 case in Bogotá, Colombia. PLoS One 16(2):e0245787.

44. Mina MJ, Parker R, & Larremore DB (2020) Rethinking Covid-19 Test Sensitivity — A Strategy for Containment. N. Engl. J. Med.

45. Ke R, et al. (2022) Longitudinal analysis of SARS-CoV-2 vaccine breakthrough infections reveal limited infectious virus shedding and restricted tissue distribution. Open Forum Infectious Diseases:ofac192.

46. Kissler SM, et al. (2021) Viral dynamics of acute SARS-CoV-2 infection and applications to diagnostic and public health strategies. PLoS Biol. 19(7):e3001333–e3001333.

47. Kissler SM, et al. (2021) Viral Dynamics of SARS-CoV-2 Variants in Vaccinated and Unvaccinated Persons. N. Engl. J. Med. 385(26):2489–2491.

48. Stankiewicz Karita HC, et al. (2022) Trajectory of Viral RNA Load Among Persons With Incident SARS-CoV-2 G614 Infection (Wuhan Strain) in Association With COVID-19 Symptom Onset and Severity. JAMA Network Open 5(1):e2142796–e2142796.

49. Deerain J, et al. (2022) Assessment of the Analytical Sensitivity of 10 Lateral Flow Devices against the SARS-CoV-2 Omicron Variant. J. Clin. Microbiol. 60(2):e0247921–e0247921.

50. Corman VM, et al. (2021) Comparison of seven commercial SARS-CoV-2 rapid point-of-care antigen tests: a single-centre laboratory evaluation study. The Lancet Microbe 2(7):e311–e319.

51. Winnett AV, et al. (2022) Morning SARS-CoV-2 testing yields better detection of infection due to higher viral loads in saliva and nasal swabs upon waking. Microbiology Spectrum.

52. Viloria Winnett A, et al. (2022) Extreme Differences in SARS-CoV-2 Omicron Viral Loads Among Specimen Types Drives Poor Performance of Nasal Rapid Antigen Tests for Detecting Presumably Pre-Infectious and Infectious Individuals, Predicting Improved Performance of Combination Specimen Antigen Tests. medRxiv:MEDRXIV/2022/277513.

53. Zymo Research (2021) SafeCollect Saliva Collection Kit User Instruction Manual. https://files.zymoresearch.com/protocols/r1211e-dna_rna_shield_safecollect_saliva_collection_kit_user_instructions.pdf

54. Zymo Research (2021) SafeCollect Swab Collection Kit User Instruction Manual. https://files.zymoresearch.com/protocols/r1160_r1161-dna_rna_shield_safecollect_swab_collection_kit_user_Instructions.pdf

55. Zymo Research (2021) Quick SARS-CoV-2 rRT-PCR Kit. https://www.zymoresearch.com/products/quick-sars-cov-2-rrt-pcr-kit

56. Cheng H-Y, et al. (2020) Contact Tracing Assessment of COVID-19 Transmission Dynamics in Taiwan and Risk at Different Exposure Periods Before and After Symptom Onset. JAMA Internal Medicine 180(9):1156–1163.

57. Rhee C, Kanjilal S, Baker M, & Klompas M (2021) Duration of Severe Acute Respiratory Syndrome Coronavirus 2 (SARS-CoV-2) Infectivity: When Is It Safe to Discontinue Isolation? Clin. Infect. Dis. 72(8):1467–1474.

58. CDC (2022) What We Know About Quarantine and Isolation. ed Centers for Disease C, and Prevention. https://www.cdc.gov/coronavirus/2019-ncov/if-you-are-sick/quarantine-isolation-background.html

59. Virtanen P, et al. (2020) SciPy 1.0: fundamental algorithms for scientific computing in Python. Nat. Methods 17(3):261–272.

60. CLSI (2008) EP12-A2 User Protocol for Evaluation of Qualitative Test Performance, 2nd edition.

61. Levy JM, et al. (2021) Impact of repeated nasal sampling on detection and quantification of SARS-CoV-2. Sci. Rep. 11(1):14903.

62. Boucau J, et al. (2022) Duration of viable virus shedding in SARS-CoV-2 omicron variant infection. medRxiv:2022.2003.2001.22271582.

63. Boucau J, et al. (2022) Duration of Shedding of Culturable Virus in SARS-CoV-2 Omicron (BA.1) Infection. N. Engl. J. Med.

64. Pray IW, et al. (2021) Performance of an Antigen-Based Test for Asymptomatic and Symptomatic SARS-CoV-2 Testing at Two University Campuses - Wisconsin, September-October 2020. MMWR Morb. Mortal. Wkly. Rep. 69(5152):1642–1647.

65. Cosimi LA, et al. (2022) Duration of Symptoms and Association With Positive Home Rapid Antigen Test Results After Infection With SARS-CoV-2. JAMA Network Open 5(8):e2225331–e2225331.

66. Bullard J, et al. (2020) Predicting Infectious Severe Acute Respiratory Syndrome Coronavirus 2 From Diagnostic Samples. Clin. Infect. Dis. 71(10):2663–2666.

67. Tom MR & Mina MJ (2020) To Interpret the SARS-CoV-2 Test, Consider the Cycle Threshold Value. Clin. Infect. Dis. 71(16):2252–2254.

68. Chu VT, et al. (2022) Comparison of Home Antigen Testing With RT-PCR and Viral Culture During the Course of SARS-CoV-2 Infection. JAMA Internal Medicine 182(7):701–709.

69. Bays D, et al. (2021) Mitigating isolation: The use of rapid antigen testing to reduce the impact of self-isolation periods. medRxiv:2021.2012.2023.21268326.

70. Pekosz A, et al. (2021) Antigen-Based Testing but Not Real-Time Polymerase Chain Reaction Correlates With Severe Acute Respiratory Syndrome Coronavirus 2 Viral Culture. Clin. Infect. Dis. 73(9):e2861–e2866.

71. Prince-Guerra JL, et al. (2021) Evaluation of Abbott BinaxNOW Rapid Antigen Test for SARS-CoV-2 Infection at Two Community-Based Testing Sites - Pima County, Arizona, November 3-17, 2020. MMWR Morb. Mortal. Wkly. Rep. 70(3):100–105.

72. Kirby JE, et al. (2022) Sars-Cov-2 antigen tests predict infectivity based on viral culture: comparison of antigen, PCR viral load, and viral culture testing on a large sample cohort. Clin. Microbiol. Infect.

73. Smith RL, et al. (2021) Longitudinal Assessment of Diagnostic Test Performance Over the Course of Acute SARS-CoV-2 Infection. The Journal of Infectious Diseases 224(6):976–982.

74. Connor BA, et al. (2022) Comparative Effectiveness of Single vs Repeated Rapid SARS-CoV-2 Antigen Testing Among Asymptomatic Individuals in a Workplace Setting. JAMA Network Open 5(3):e223073–e223073.

75. Korenkov M, et al. (2021) Evaluation of a Rapid Antigen Test To Detect SARS-CoV-2 Infection and Identify Potentially Infectious Individuals. J. Clin. Microbiol. 59(9):e00896–00821.

76. Wyllie AL, et al. (2020) Saliva is more sensitive for SARS-CoV-2 detection in COVID-19 patients than nasopharyngeal swabs. medRxiv:2020.2004.2016.20067835.

77. Thomas HM, et al. (2022) Acceptability of OP/Na swabbing for SARS-CoV-2: a prospective observational cohort surveillance study in Western Australian schools. BMJ Open 12(1):e055217.

78. National Health Service (2021) How to use an NHS rapid lateral flow test for coronavirus (COVID-19). https://www.nhs.uk/conditions/coronavirus-covid-19/testing/how-to-do-a-test-at-home-or-at-a-test-site/how-to-do-a-rapid-lateral-flow-test/

79. National Health Service (2021) How to use an NHS PCR test for coronavirus (COVID-19). https://www.nhs.uk/conditions/coronavirus-covid-19/testing/how-to-do-a-test-at-home-or-at-a-test-site/how-to-do-a-pcr-test/

80. FDA (2022) In Vitro Diagnostic EUAs. https://www.fda.gov/medical-devices/coronavirus-disease-2019-covid-19-emergency-use-authorizations-medical-devices/in-vitro-diagnostics-euas

81. LeBlanc JJ, et al. (2020) A combined oropharyngeal/nares swab is a suitable alternative to nasopharyngeal swabs for the detection of SARS-CoV-2. J. Clin. Virol. 128:104442.

82. Vlek ALM, Wesselius TS, Achterberg R, & Thijsen SFT (2021) Combined throat/nasal swab sampling for SARS-CoV-2 is equivalent to nasopharyngeal sampling. European journal of clinical microbiology & infectious diseases: official publication of the European Society of Clinical Microbiology 40(1):193–195.

83. Lee Rose A, et al. (Performance of Saliva, Oropharyngeal Swabs, and Nasal Swabs for SARS-CoV-2 Molecular Detection: a Systematic Review and Meta-analysis. J. Clin. Microbiol. 59(5):e02881–02820.

84. Shakir SM, et al. (2020) Combined Self-Collected Anterior Nasal and Oropharyngeal Specimens versus Provider-Collected Nasopharyngeal Swabs for the Detection of SARS-CoV-2. J. Clin. Microbiol. 59(1).

85. Desmet T, et al. (2021) Combined oropharyngeal/nasal swab is equivalent to nasopharyngeal sampling for SARS-CoV-2 diagnostic PCR. BMC Microbiol. 21(1):31.

86. Pan Y, Zhang D, Yang P, Poon LLM, & Wang Q (2020) Viral load of SARS-CoV-2 in clinical samples. The Lancet Infectious Diseases 20(4):411–412.

87. Yan Y, Chang L, & Wang L (2020) Laboratory testing of SARS-CoV, MERS-CoV, and SARS-CoV-2 (2019-nCoV): Current status, challenges, and countermeasures. Rev. Med. Virol. 30(3):e2106.

88. Mawaddah A, Gendeh HS, Lum SG, & Marina MB (2020) Upper respiratory tract sampling in COVID-19. Malays. J. Pathol. 42(1):23–35.

89. Manabe YC, et al. (2021) Self-Collected Oral Fluid Saliva Is Insensitive Compared With Nasal-Oropharyngeal Swabs in the Detection of Severe Acute Respiratory Syndrome Coronavirus 2 in Outpatients. Open Forum Infectious Diseases 8(2).

90. Sharma K, et al. (2021) Comparative analysis of various clinical specimens in detection of SARS-CoV-2 using rRT-PCR in new and follow up cases of COVID-19 infection: Quest for the best choice. PLoS One 16(4):e0249408.

91. Michel W, et al. (2021) A combined oro-nasopharyngeal swab is more sensitive than mouthwash in detecting SARS-CoV-2 by a high-throughput PCR assay. Infection 49(3):527–531.

92. Jeong HW, et al. (2020) Viable SARS-CoV-2 in various specimens from COVID-19 patients. Clin. Microbiol. Infect. 26(11):1520–1524.

